# Number of Trials and E-Field Orientation during Continuous Theta Burst Stimulation May Impact Modulation of Motor-Evoked Potentials

**DOI:** 10.1101/2024.05.23.24307821

**Authors:** Silas Preis, Su Hwan Kim, Paul Schandelmaier, Claus Zimmer, Bernhard Meyer, Sandro M. Krieg, Nico Sollmann, Severin Schramm

## Abstract

**Introduction:** Noninvasive neuromodulation (NM) via transcranial magnetic stimulation (TMS) is increasingly applied to treat neurological and psychiatric disorders. However, NM effects are highly variable between subjects. E-field orientation (EFO) during NM protocols may heavily contribute to this variability. Investigating the influence of EFO during NM could lead to improved therapeutic protocols by enabling more tailored approaches for patient-specific NM. In the present study, we aimed to examine the influence of varying EFO during continuous theta burst stimulation (cTBS) on the modulation of motor-evoked potentials (MEPs).

**Methods:** 20 healthy volunteers (8 F; mean age 25.7±2.7 years) took part in this prospective, single blind sham-controlled crossover study consisting of three neuronavigated TMS sessions. The sessions differed only in EFO during cTBS (parallel to optimal EFO for MEP generation [OPT], 90° rotated from OPT [90], 45° rotated from OPT with 7.3 cm spacer [SHAM]). Electromyography was recorded from abductor pollicis brevis, first dorsal interosseous, and adductor digiti minimi muscles during stimulation of the abductor pollicis brevis (APB) motor hotspot. 4 blocks (PRE, POST1, POST2, POST3) with 30 MEPs each were elicited from the motor hotspot. Between the PRE and POST1 block, 40 s of cTBS were performed using one of the three EFO paradigms. Individual POST blocks were separated by a 2 min interval. MEPs were analyzed with linear mixed effects modeling augmented by bootstrapping.

**Results:** A total of 19,830 MEPs were analyzed. Progression through the trial blocks led to heightened MEP amplitudes (e.g., POST3 vs. PRE; log-estimate 0.244, t = 21.43), and later trials were significantly associated with higher MEP amplitudes (spearman’s rho 0.981; p < 0.001). Additionally, on the group level, a significant albeit slight influence of EFO on MEP amplitudes with the 90 paradigm leading to facilitation, and SHAM paradigm leading to suppression of MEP amplitudes was observed when compared to the OPT paradigm (log-estimate 90: 0.135, t = 13.604; log-estimate SHAM: −0.043, t = −4.283). On the subject level, we observed strong heterogeneity between individuals regarding their response to cTBS using varying EFO.

**Discussion:** We observed that MEP amplitudes following cTBS differed significantly based on EFO during NM. This implies that for a given desired NM result, individual EFO optimization may act as an avenue to maximize the NM effect. Therapeutic NM applications might consider EFO as a parameter of interest to be investigated in clinical studies. Additionally, prolonged single-pulse stimulation appeared to possess a NM quality of its own, which should be considered in TMS studies employing single-pulse protocols.

## 1. Introduction

Transcranial magnetic stimulation (TMS) is a method that enables noninvasive stimulation of the cerebral cortex for a variety of use cases. In recent years, neuromodulation (NM) via TMS has seen increasing adoption in different clinical applications ^1–4^. While the empirical therapeutic benefits of TMS-based NM are, depending on the specific diagnosis to be treated, quite considerable on the group level, recent work has proposed that NM responses are subject to notable heterogeneity ^5–10^. Sources for these heterogeneous responses can be found in a variety of factors, including but not limited to brain state as assessed via electroencephalography (EEG) ^7, 11^, individual functional connectivity as assed via functional magnetic resonance imaging (fMRI) ^12^, targeting the region of interest via external anatomical structures or via neuronavigation ^13^, stimulation target selection ^14^, and pulse shape ^15^.

Another factor relevant in TMS is the e-field orientation (EFO). During TMS, the induced e-field interacts with underlying gyri and the neuron populations contained therein ^16^. Varying the local EFO by e.g. rotating the TMS coil has been demonstrated to influence both the distribution of the cortical electric field as well as outcome parameters such as EEG-derived changes or motor-evoked potentials (MEPs) elicited in single-pulse protocols ^17–22^. Aside from the shape and strength of the electric field itself, the heterogeneous shape and occurrence of different neuron populations across cortical layers contribute to the differential sensitivities regarding TMS applied at varying EFO ^23^. From this follows that EFO may plausibly influence results of NM by TMS.

However, to date, a systematic evaluation of the influence of EFO regarding the effects of NM protocols on MEPs has not yet been performed in neuronavigated TMS. Therefore, we aimed to investigate whether variations in EFO during continuous theta burst stimulation (cTBS), a commonly employed protocol for MEP modulation ^24–26^, could cause differential modulation of subsequent MEPs.

## 2. Methods

### 2.1. Ethics and study design

Approval for the presented study was granted by the local institutional review board (Ethics Committee of Technical University of Munich). The study was conducted in accordance with the Declaration of Helsinki. All subjects declared written informed consent.

The present study was conceptualized as a prospective cross-over single-blind sham-controlled study. A power analysis was conducted with the following assumptions: level of statistical significance 0.05; standard deviation of MEP amplitudes 400 µV (taken from a comparable study ^9^); power 0.95; minimal detectable difference in means 500 µV; yielding a need for 19 participants. Due to one exclusion after their first session (see results), 20 different subjects were recruited.

### 2.2. Participant selection

Healthy participants were recruited via word of mouth. Inclusion criteria were defined as: age > 18 years; written informed consent. Exclusion criteria were defined as: any history of neurological or psychiatric diseases; incidental findings on cerebral MRI (e.g., intracranial aneurysm); any contraindications for MRI or TMS (e.g., implanted ferromagnetic devices, pregnancy).

### 2.3. Structural imaging for neuronavigation

Before taking part in the TMS experiment, participants underwent an MRI protocol containing a three-dimensional (3D) T1-weighted gradient echo sequence (repetition time/echo time: 9/4 ms, 1 mm^3^ isovoxel covering the whole head; 3-T Philips Ingenia, Philips Healthcare, Best, The Netherlands). This imaging sequence was used for subsequent neuronavigation during TMS. All images acquired were examined for incidental findings by at least two physicians from the neuroradiology department.

### 2.4. TMS experiment

Stimulation by TMS was conducted using the Nexstim NBS system (Nexstim eXimia NBS system, version 5.1; Nexstim Plc., Helsinki, Finland). The system allows for neuronavigation based on reverse coregistration of the patient to the structural imaging and employs a biphasic pulse form for stimulation ^27, 28^.

Each participant underwent three TMS sessions in randomized order differing only regarding EFO during NM (Figure 1). Any randomizations were conducted via the “Sequence Generator” function of the homepage www.random.org. Sessions were spaced at least 14 days apart from another to avoid carry-over effects from previous sessions. Electromyography (EMG) recordings were conducted using surface electrodes (Neuroline 720; Ambu, Ballerup, Denmark) for three muscles on the dominant hand of the participant: abductor pollicis brevis (APB), adductor digiti minimi (ADM), and first dorsal interosseous (FDI) muscles. For MEP generation, the inter-pulse interval was randomized to 5.4 - 6.6 s.

**Figure 1:**
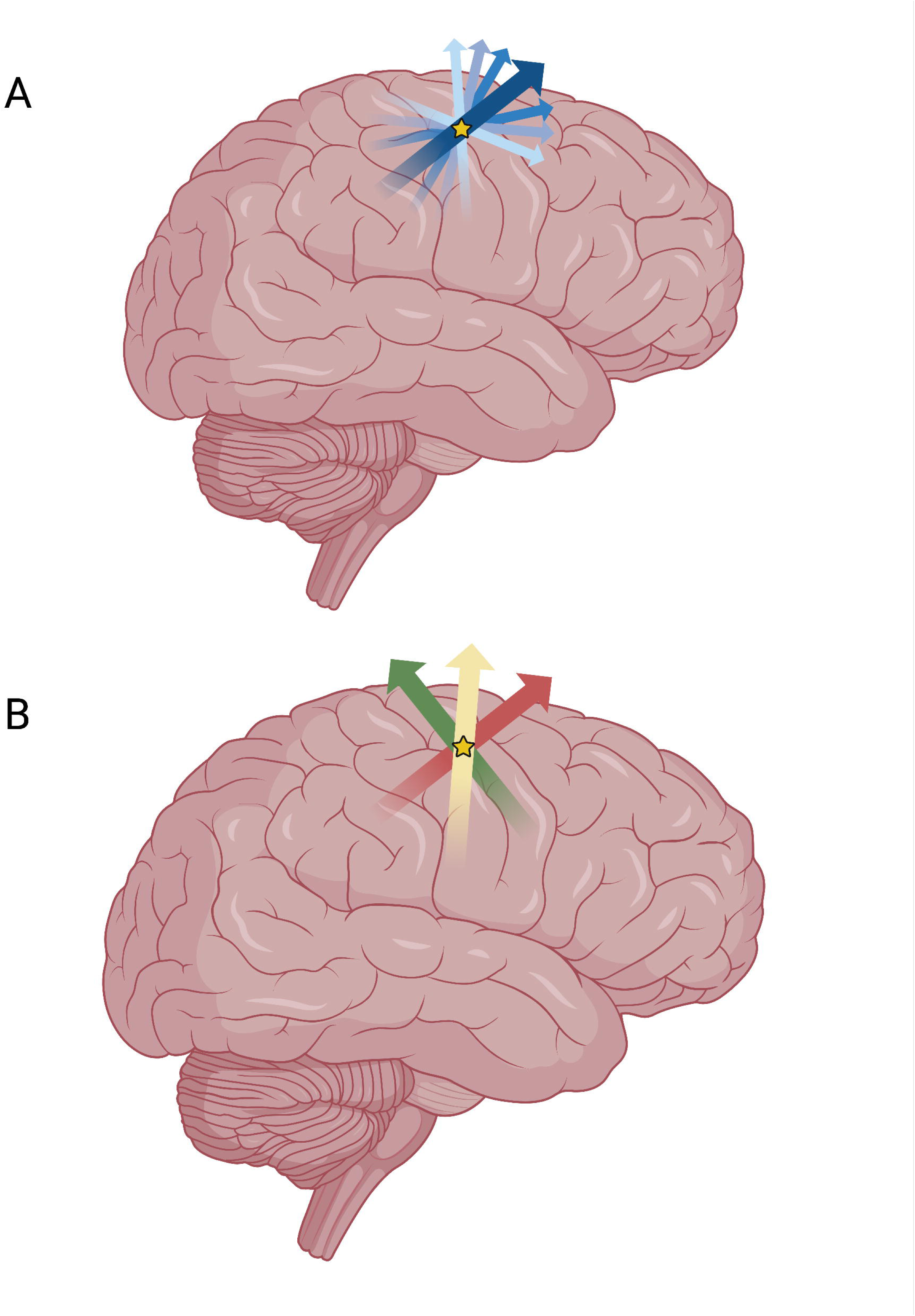
Determination of Theta Burst Stimulation Directions. Figure 1 demonstrates the process by which the directional conditions (OPT, 90, SHAM) for continuous theta burst stimulation (cTBS) were determined. A) In the beginning of each session, a main direction (large blue arrow) was defined based on a 90° orientation compared to the course of the precentral gyrus. Based on this, 7 directions separated by 20° steps were stimulated (all blue arrows) over the abductor pollicis brevis motor hotspot (star) in randomized order. B) The direction with maximum mean MEP amplitude was chosen to represent OPT (red arrow). The direction 90 was chosen by rotating the coil 90° away from OPT (green arrow). While no effective stimulation was conducted during the SHAM paradigm (performed with 7.3 cm spacer), the directionality was chosen as the midway between OPT and 90 (yellow arrow).

#### 2.4.1. Determination of APB motor hotspot and EFO conditions

During the first session, the APB motor hotspot on the hemisphere contralateral to the dominant hand (e. g. left hemisphere for right handed individuals) was determined according to clinical practice ^29^ ^28^. Since MEP generation is sensitive to current direction ^30, 31^, the optimal EFO for MEP generation was determined by rotating the coil across 7 different directions in randomized order (Figure 1). In this context, the reference direction was defined as 90° in relation to the course of the precentral gyrus, while the additional directions were defined as being ± 20°, ± 40° and ± 60° rotated from the reference direction, yielding 7 directions in total (Figure 1). The sequence in which these directions were tested was randomized for each appointment. In total, 20 MEPs were elicited for each EFO tested this way. The intensity during this step was selected so that MEPs in the reference direction were approximately in the 200-500 µV range. The intensity setting at this stage was not based on resting motor threshold (rMT), since at this point in the experiment, the optimal direction for rMT determination was not yet known.

The EFO yielding the highest mean MEP amplitude (average of the 20 MEPs per direction) was considered the hypothetically optimal EFO (“OPT”) in the frame of subsequent cTBS application. The alternative condition “90” was defined by rotation of the coil 90° away from OPT (clockwise in reference to left hemisphere, counter-clockwise in reference to right hemisphere; Figure 1). The third condition “SHAM” was realized by introducing a spacer between the coil and the scalp, adding 7.3 cm distance and thus prohibiting effective stimulation while retaining the sensory feeling of a coil as well as the sounds of the stimulation. Additionally, the coil was rotated 45° away from OPT during SHAM (Figure 1).

The EFO for OPT, 90, and SHAM paradigms were kept constant as defined during this first session across the subsequent sessions. To avoid bias to the data by varying the total number of stimuli, the 20 MEPs across 7 directions were also elicited in the subsequent sessions, but not again considered for EFO determination.

#### 2.4.2. MEP generation and cTBS application

After determination of OPT, the rMT (in maximum stimulator output [MSO]) was measured in each session according to the maximum-likelihood algorithm ^28, 32^. Afterwards, the first of four MEP generation blocks took place (Figure 2). In the PRE-block, 30 MEPs were elicited in OPT direction at 150% rMT ^33, 34^. Subsequently, cTBS in either OPT, 90, or SHAM condition was performed according to the classic paradigm by Huang et al., consisting of 3 pulses at 50 Hz every 200 ms, for 600 pulses in total over 40 s ^26^. The cTBS paradigm was applied at 70% rMT as in comparable studies ^33, 35^. Immediately afterwards, the rMT was redetermined. This was then followed by another 30 MEPs at 150% of the initial rMT (POST1), again in OPT EFO. After 2 min, the POST2 block was conducted in a similar manner, followed by POST3 again 2 min later. The total number of pulses thus applied over the 4 blocks was 120 (excluding pulses during rMT determination). An overview of the experimental procedure is given in Figure 2.

**Figure 2:**
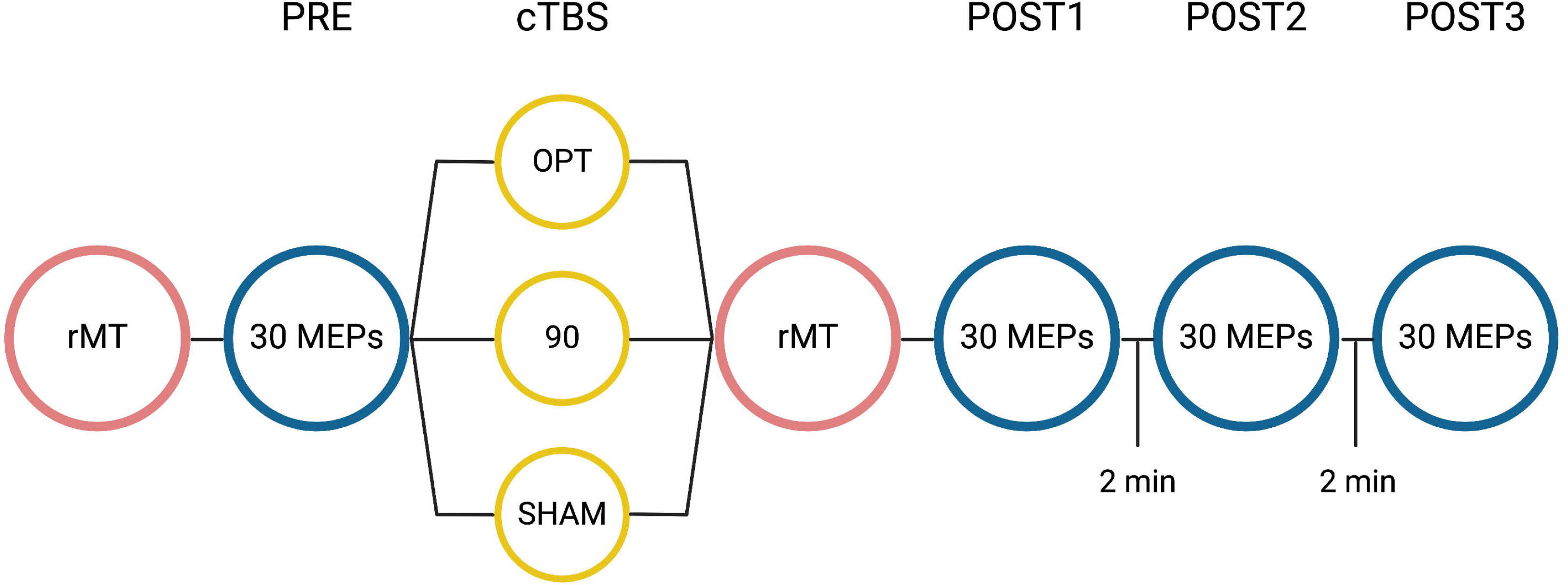
Schematic Display of Experiment Design. Figure 2 demonstrates the experimental design after determination of OPT, 90 and SHAM (Figure 1). After rMT determination in OPT direction, the first block (PRE) of 30 motor evoked potentials (MEPs) was performed at 150 % rMT. Afterwards, one of three continuous theta burst (cTBS) paradigms was performed at 70 % rMT, followed by immediate redetermination of rMT. Afterwards, three blocks of 30 MEPs each were conducted, interspersed by 2 minutes each for a total interval of 5 minutes from beginning of one block to the next.

After data generation, the automatic MEP amplitude measurements conducted by the NBS system were inspected and corrected when necessary, e. g. when MEP onsets were misidentified or peak-to-peak amplitude was measured incorrectly. Specifically, MEPs were considered valid if their amplitude was > 50 µV and the latency was plausible (i.e., 15- 30 ms for stimulation of upper extremity muscles ^36, 37^). The corrected MEPs were exported to an excel file.

### 2.5. Statistical analysis

Statistical analyses took place in RStudio (RStudio, 2022.02.3 Build 492; © 2009-2022 RStudio, PBC). Since initial data inspection revealed an apparent trend across all cTBS paradigms towards higher MEPs, an analysis of variance (ANOVA) and subsequent Spearman correlation analysis were performed to investigate the correlation between trial number (1-120) and MEP amplitude. For analysis of the impact of the cTBS paradigm on MEP amplitude, an ANOVA with post-hoc testing by TukeyHSD test was performed.

A mixed effects linear regression model was employed to analyze the effect of our theory-motivated (cTBS condition) as well as our data-driven (trial block) explanatory variables on our response variable (MEP amplitude). These were modeled as fixed effects. Additionally, we included the stimulation intensity in the model as a fixed effect to account for its potential effect on MEP amplitude. Lastly, we modeled a random intercept for each interaction between subject and muscle, to account for idiosyncrasies of each subject and potential muscle-related variations across subjects. All modeling was done using the lme4 package ^38^.

An initial attempt at modeling suggested significant heteroscedasticity within the data (supplementary material). Therefore, the final modeling was conducted after applying a natural logarithmic transform to all MEP amplitudes. The coefficients of the final model were further investigated by employing the bootstrapping method (as implemented in the bootMer function of the lme4 package ^38^) to fit the same model to 10,000 resampled datasets and observe distributions of both the coefficients as well as the corresponding t-values (Table 3). A p-value < 0.05 was considered statistically significant.

## 3. Results

Out of 20 recruited subjects (8 f; mean age 25.7 ± 2.7 years), one subject was excluded after the initial session due to an incidental finding on MRI (cerebral aneurysm). The data gained from their first session, which had taken place in the interim, was still used in the subsequent analysis.

Overall, 19,830 MEPs were acquired and included in the statistical analysis. During 5 appointments, no recording of the FDI could be acquired, while for the ADM, no recordings could be acquired during 3 appointments (see subject-specific data in supplementary information). Additionally, in one subject, the POST3 block was not acquired in one session due to patient discomfort. No significant differences in rMT before and after cTBS based on EFO paradigm were detected (overall rMT before cTBS: 33 ± 7% MSO; rMT after cTBS: 34 ± 7% MSO). Descriptive statistics on MEPs and rMTs split by muscle and cTBS condition can be found in Table 1.

**Table 1:**
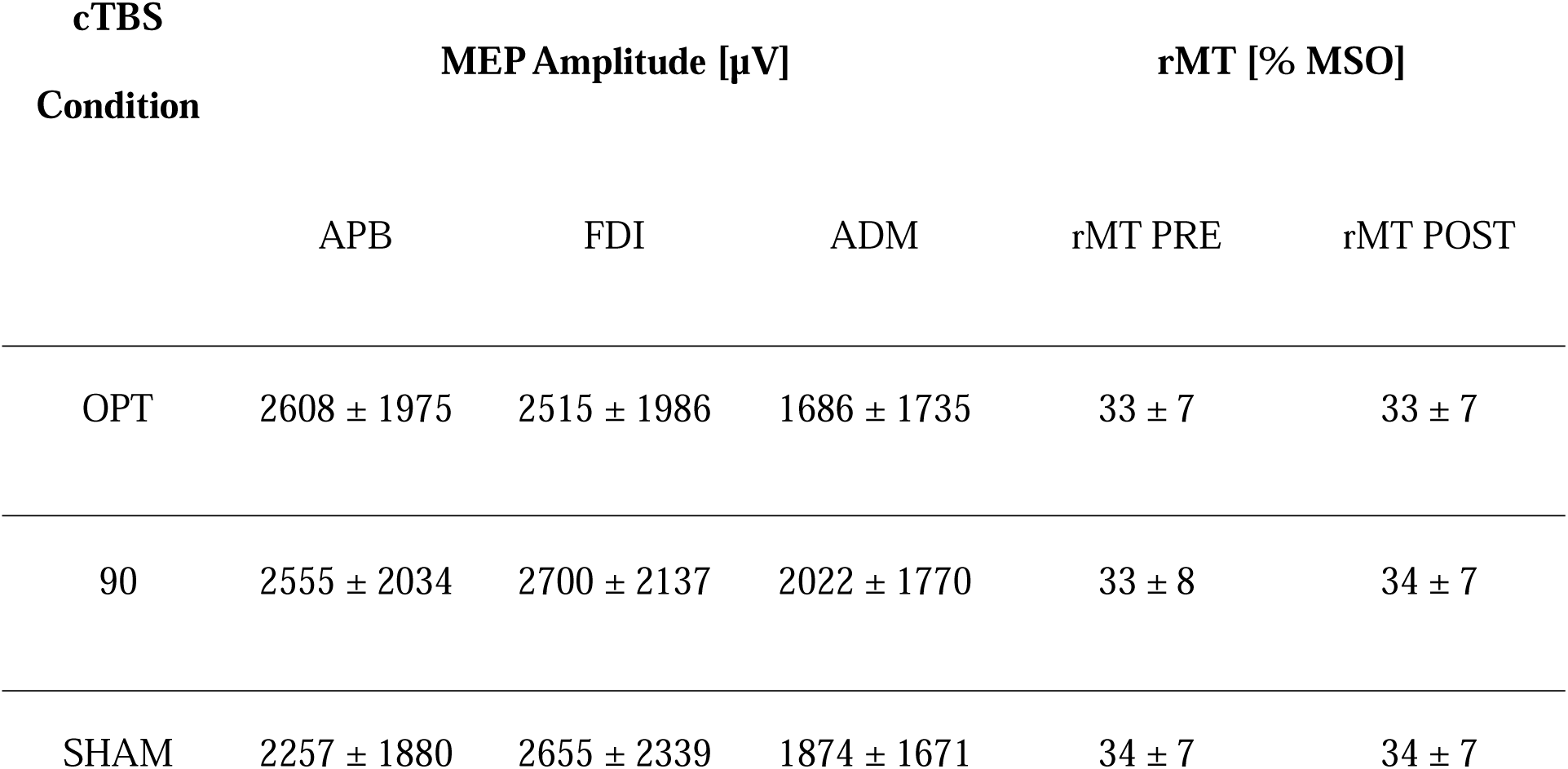
Descriptive statistics of MEP amplitudes across all participants. Table 1 yields an overview over all collected motor evoked potential (MEP) amplitudes and resting motor thresholds (rMT) split according to session-dependent cTBS condition (OPT, 90, SHAM) and muscle. Values are given in µV and in the format of “mean ± standard deviation”. Abbreviations: Abductor pollicis brevis (APB), adductor digiti minimi (ADM), first dorsal interosseous (FDI), maximum stimulator output (MSO), resting motor threshold (rMT).

| cTBS<br>Condition | MEP Amplitude [μV] | | | rMT [% MSO] | |
| --- | --- | --- | --- | --- | --- |
|  | APB | FDI | ADM | rMT PRE | rMT POST |
| OPT | 2608 ± 1975 | 2515 ± 1986 | 1686 ± 1735 | 33 ± 7 | 33 ± 7 |
| 90 | 2555 ± 2034 | 2700 ± 2137 | 2022 ± 1770 | 33 ± 8 | 34 ± 7 |
| SHAM | 2257 ± 1880 | 2655 ± 2339 | 1874 ± 1671 | 34 ± 7 | 34 ± 7 |

### 3.1. Effect of trial number

Initial data inspection indicated a trend of rising MEP amplitudes shared by all muscles and cTBS conditions (Figure 3A-C), which was confirmed by ANOVA (p < 0.0001). Notably, the trend also appeared to be present in the SHAM condition (Figure 3B). Additionally, in the group level analysis, significant differences in MEP amplitudes were not observed between PRE and POST1, and neither between POST2 and POST3. This was understood to imply a significant, albeit potentially nonlinear relationship between additional trials and higher MEP amplitude.

**Figure 3:**
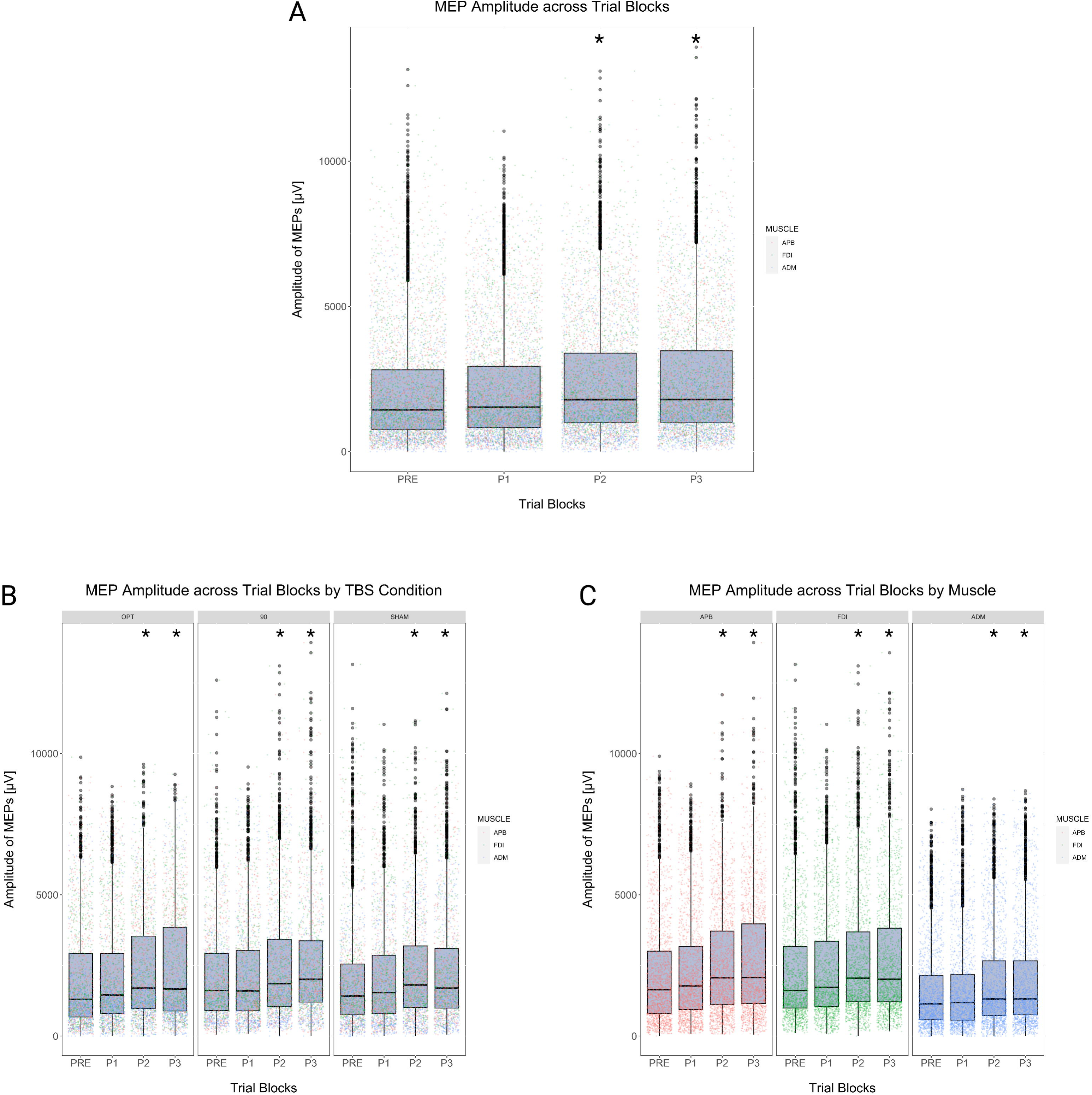
MEP Amplitudes across Trial Blocks. Figure 3 demonstrates the MEP amplitudes across trial blocks (PRE, POST1-3 [P1-3]) in total and in various subgroups. A) Boxplots of pooled MEP amplitudes across all continuous theta burst stimulation (cTBS) paradigms (OPT, 90, SHAM). Colors denote different muscles (red: abductor pollicis brevis; green: first dorsal interosseous; blue: adductor digiti minimi). B) Boxplots of MEP amplitudes split by cTBS paradigm across trial blocks. Colors denote different muscles (red: abductor pollicis brevis; green: first dorsal interosseous; blue: adductor digiti minimi). C) Boxplots of MEP amplitudes split by muscles across trial blocks. For ease of visualization, only significant differences of POST blocks compared to PRE blocks are denoted via star (p < 0.05, corrected by Tukey procedure within each subplot). Significant differences between individual POST blocks are not visualized.

Thus, a Spearman rank correlation analysis was conducted to determine whether the position in the trial sequence (e. g. 1^st^ MEP – 120^th^ MEP) correlated with MEP amplitude. The corresponding test revealed a slight, yet highly significant correlation (Spearman’s rho 0.0981; p < 0.0001; Figure 4).

**Figure 4:**
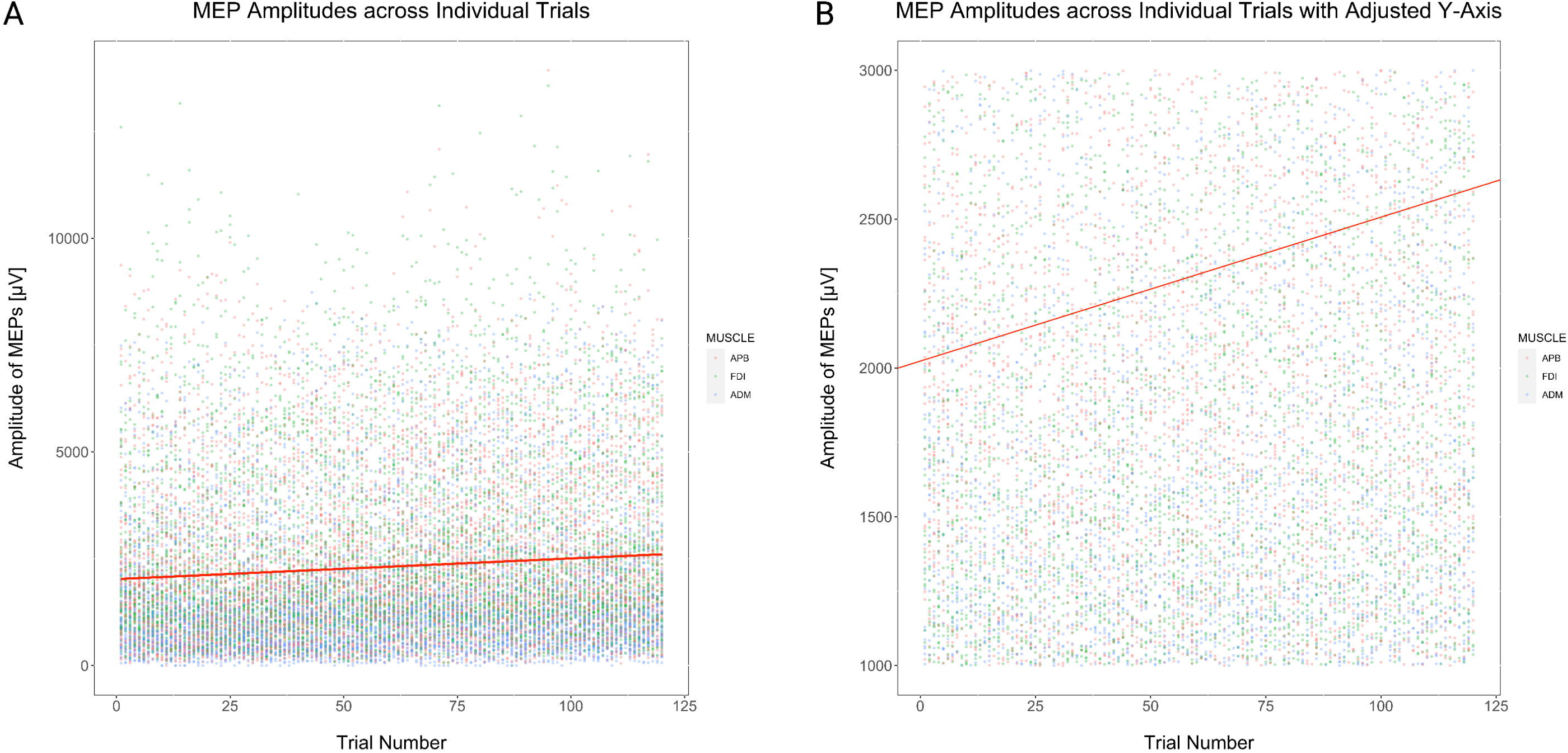
MEP Amplitudes across Individual Trials. Figure 4 demonstrates the MEP amplitudes across individual trials from all trial blocks (trials 1-120). A) Scatterplot of pooled MEP amplitudes across all continuous theta burst stimulation (cTBS) paradigms (OPT, 90, SHAM). Colors denote different muscles (red: abductor pollicis brevis; green: first dorsal interosseous; blue: adductor digiti minimi). The red line indicates a linear regression line visualizing the correlation between MEP amplitudes and trial number. B) Analogous plot to A) with adjusted y-axis for better visualization of regression line.

We thus decided to incorporate the information on trial block into our subsequent mixed effects regression model. In the model, each subsequent trial block was estimated to have a significant facilitating effect on MEP amplitudes compared to the PRE timepoint (log-estimate POST1: 0.058; t = 5.135 / POST2: 0.208; t = 18.385 / POST: 0.244; t = 21.431; Table 2). The bootstrapped estimates indicated no differences regarding statistical significance from the original estimates with only minor biases (Table 3).

**Table 2:**
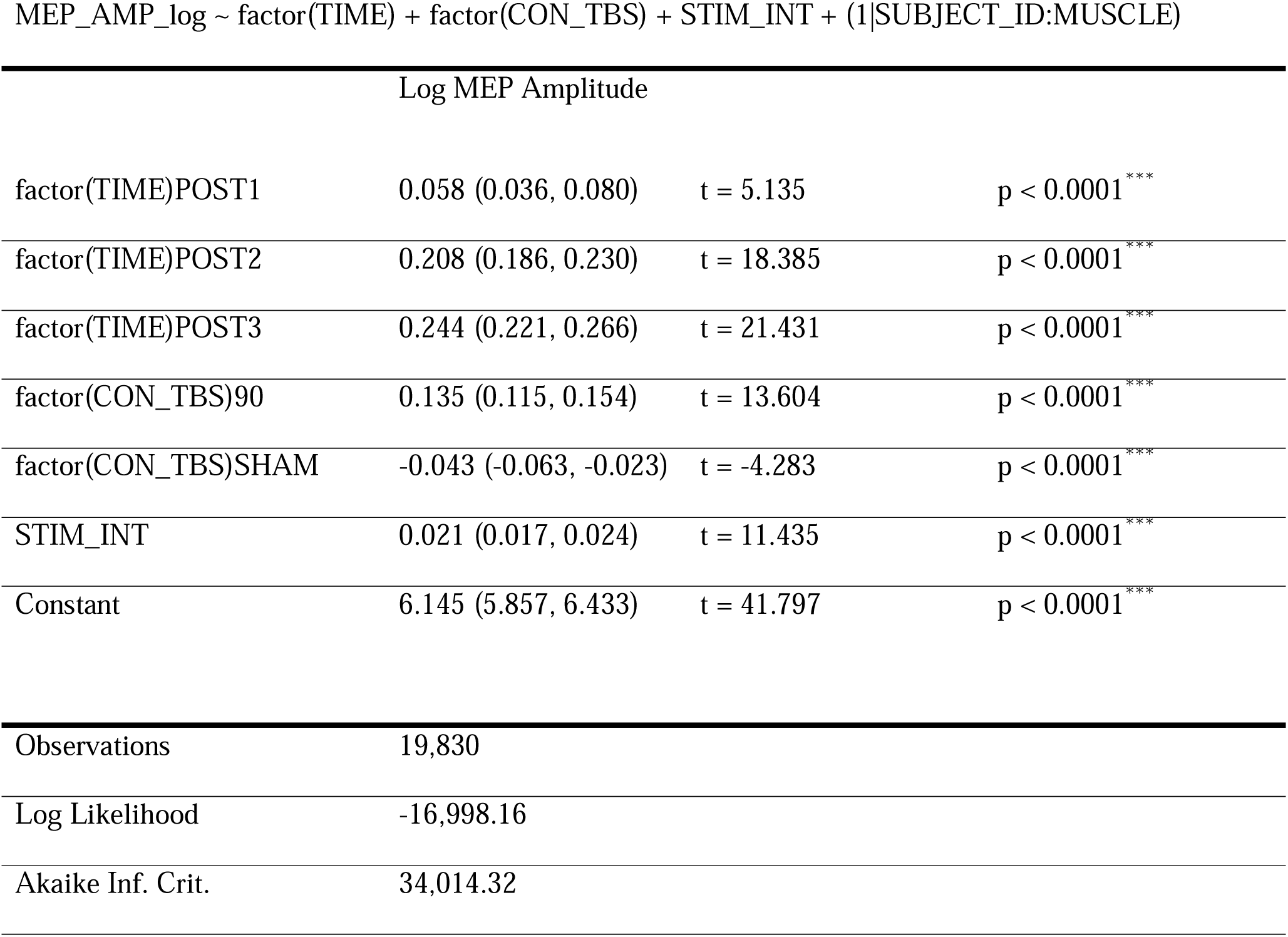
Linear mixed effects regression model. Table 2 yields an overview over the conducted mixed effects linear regression model. The model specification is given in the first line. TIME refers to a 4-level ordinal factor indicating the trial block (PRE, POST1, POST2, POST3). CON_TBS refers to a 3-level nominal indicating the theta burst condition (OPT, 90, SHAM). STIM-INT is a numeric indicating stimulation intensity in percent of maximum stimulator output. SUBJECT_ID refers to a 20-level nominal indicating the subject, MUSCLE refers to a 3-level nominal indicating the muscle from which the MEP was taken. Coefficient estimates are presented in the format of “estimate (lower confidence interval, upper confidence interval)”.

**Table 3:** Bootstrap statistics of the linear mixed effects regression model. Table 3 yields an overview over the conducted bootstrapping statistics on the model specified in Table 2 based on n(sim) = 10,000. Explanatory variables are idem to Table 2.

| **Coefficients** | Estimate | Bias | Lower CI | Upper CI |
| --- | --- | --- | --- | --- |
| (Intercept) | 6.144899 | -3.96E-04 | 5.854066 | 6.435576 |
| factor(TIME)POST1 | 0.058025 | 9.27E-05 | 0.035544 | 0.080928 |
| factor(TIME)POST2 | 0.207752 | -1.72E-04 | 0.185022 | 0.230384 |
| factor(TIME)POST3 | 0.243659 | -1.28E-04 | 0.221182 | 0.266451 |
| factor(CON_TBS)90 | 0.134797 | 1.50E-04 | 0.11548 | 0.154428 |
| factor(CON_TBS)SHAM | -0.04303 | 6.89E-06 | -0.06266 | -0.02362 |
| STIM_INT | 0.020548 | -1.10E-05 | 0.016986 | 0.024057 |
| **t-values** |  |  |  |  |
| (Intercept) | 41.79665 | 2.26E-01 | 37.19361 | 47.33439 |
| factor(TIME)POST1 | 5.134858 | 8.31E-03 | 3.145556 | 7.167981 |
| factor(TIME)POST2 | 18.3848 | -1.50E-02 | 16.37821 | 20.38887 |
| factor(TIME)POST3 | 21.43146 | -1.10E-02 | 19.41618 | 23.44726 |
| factor(CON_TBS)90 | 13.60402 | 1.54E-02 | 11.6391 | 15.57816 |
| factor(CON_TBS)SHAM | -4.28317 | 5.84E-04 | -6.24038 | -2.34912 |
| STIM_INT | 11.43518 | -1.55E-05 | 9.43797 | 13.39882 |

### 3.2. Effect of EFO during cTBS

A granular visualization of the MEP data revealed no clear differential trends split by cTBS conditions and muscles (Figure 5). ANOVAs and subsequent post-hoc testing revealed a number of significant differences between trial blocks, exclusively demonstrating MEP facilitation on the group level (Figure 5).

**Figure 5:**
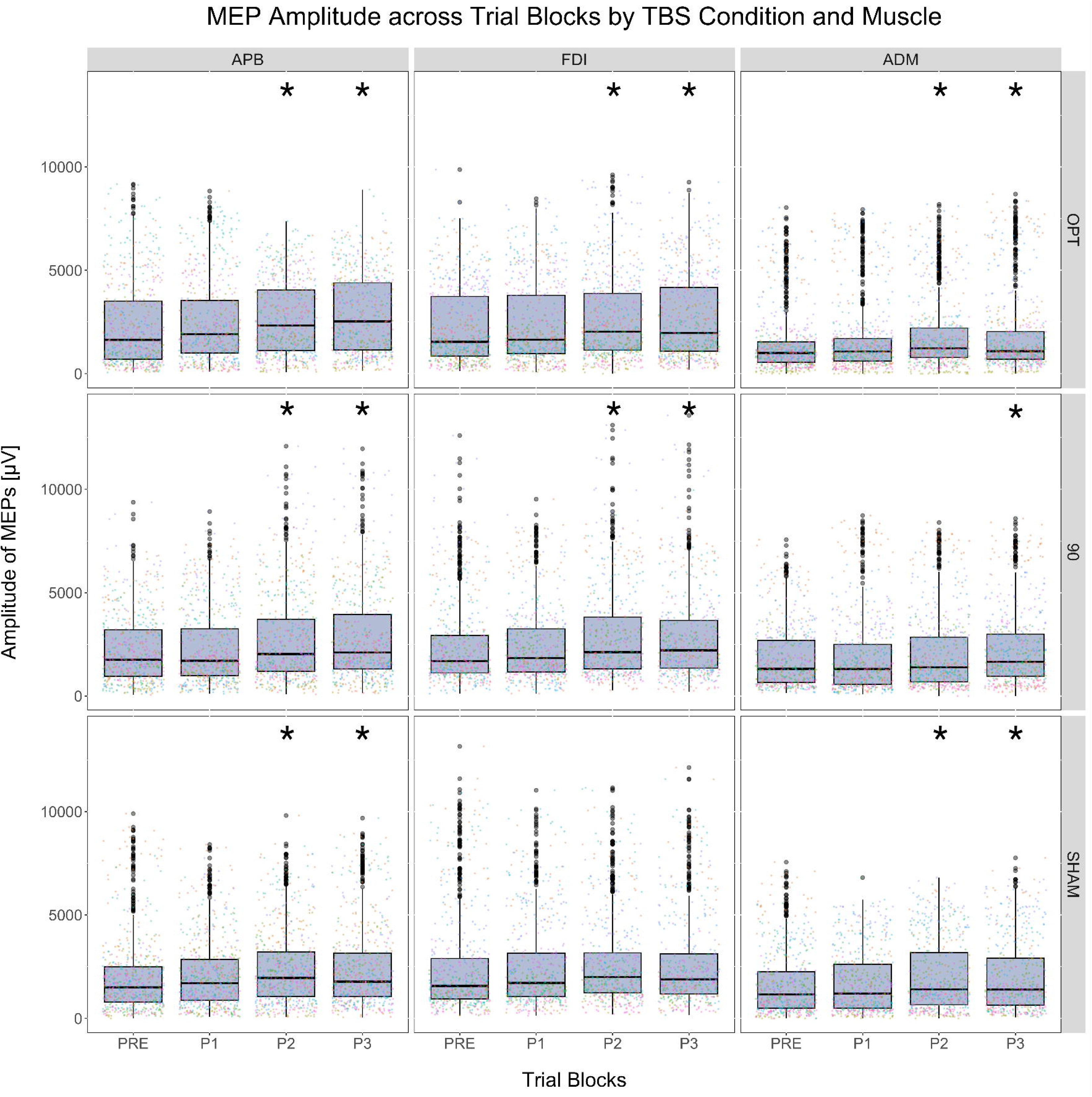
MEP Amplitudes across Trial Blocks Split by Muscle and Theta Burst Condition. Figure 5 demonstrates the MEP amplitudes across all trial blocks (PRE, POST1-3 [P1-3]) split by muscle and continuous theta burst stimulation (cTBS) paradigm. Color denotes individual subjects from which the data was recorded. For ease of visualization, only significant differences of POST blocks compared to PRE blocks are denoted via star (p < 0.05, corrected by Tukey procedure within each subplot). Significant differences between individual POST blocks are not visualized.

In the linear mixed-effects regression model, significant differences between cTBS conditions emerged (Table 2). Compared to OPT, the 90 paradigm was estimated to lead to significantly higher MEP amplitudes (log-estimate 0.135; t = 13.604), corresponding to a comparative factorial increase of e^0.135^ ≈ 1.14.

Conversely, the SHAM condition was predicted to lead to a comparative MEP decrease vice versa OPT (log-estimate −0.043; t = −4.283), corresponding to an approximate e^-0.043^ ≈ 0.95 factorial decrease of MEP amplitudes. Again, the bootstrapped estimates indicated no differences regarding statistical significance from the original estimates with only minor biases (Table 3).

### 3.3. Heterogeneity on single-subject level

When visualizing data on the single-subject level, considerable heterogeneity in response to cTBS with varying EFO was noted, with MEP facilitation occurring after OPT cTBS and suppression occurring after 90 cTBS in one case, while the exact opposite trend was observed in another case (Figure 6, subjects 6 and 7). Additionally, another subject demonstrated suppression of MEP amplitudes after SHAM cTBS contrasted by facilitation after both OPT and 90 cTBS (Figure 6, subject 2). Additional case-specific data visualizations can be found in the supplementary material.

**Figure 6:**
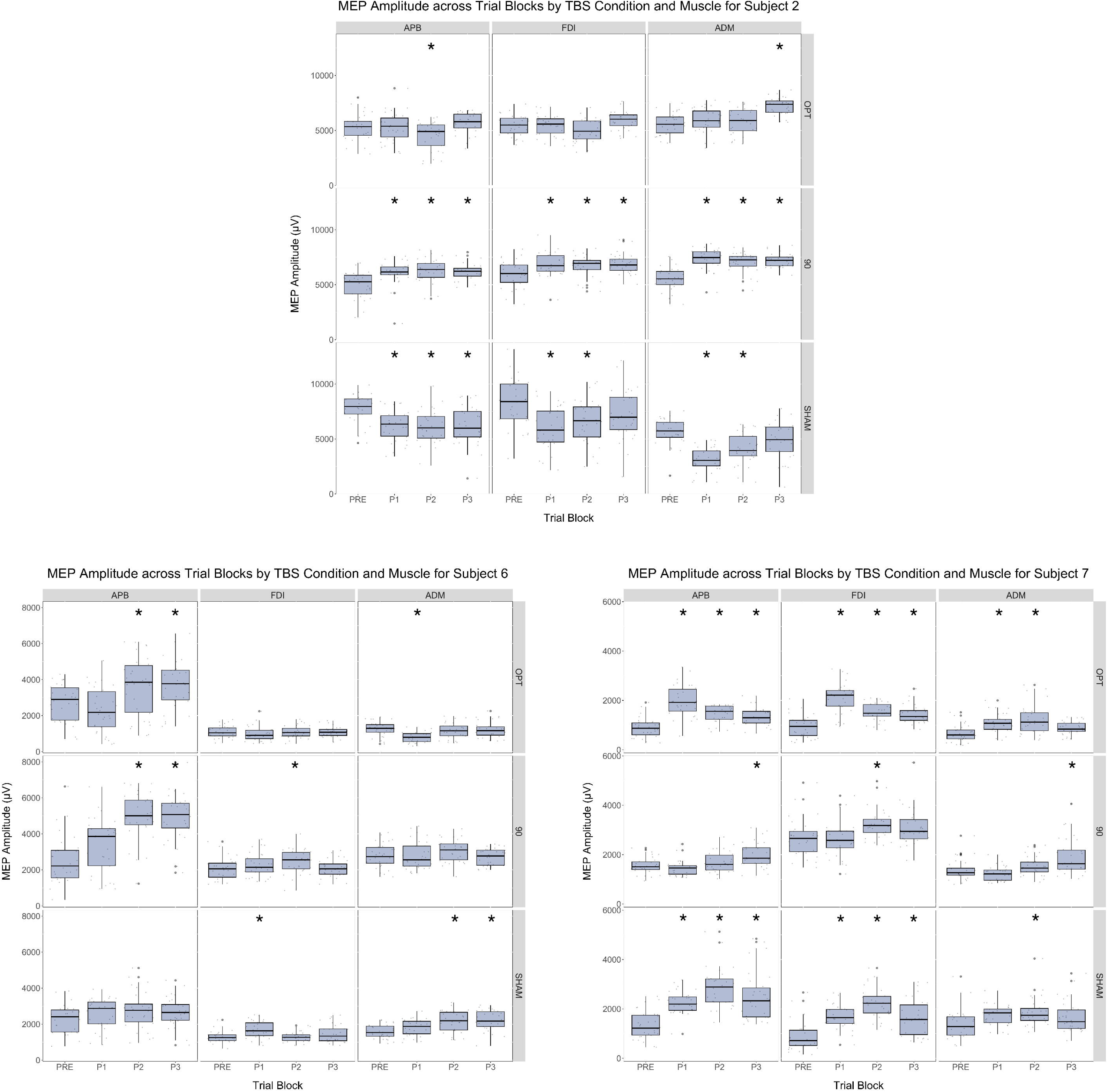
Single-subject MEP Amplitudes across Trial Blocks Split by Muscle and Theta Burst Condition. Figure 6 demonstrates the MEP amplitudes across all trial blocks (PRE, POST1-3 [P1-3]) split by muscle and continuous theta burst stimulation (cTBS) paradigm for subjects 2, 6 and 7 respectively as exemplary cases. Note the high heterogeneity in response to continuous theta burst stimulation (cTBS) paradigms across different subjects. For ease of visualization, only significant differences of POST blocks compared to PRE blocks are denoted via star (p < 0.05, corrected by Tukey procedure within each subplot). Significant differences between individual POST blocks are not visualized.

## 4. Discussion

In the present study, we assessed cTBS-based NM of MEPs in healthy subjects across two verum and one sham condition to investigate the impact of EFO during cTBS. We observed a significant, albeit slight impact of EFO on MEP amplitudes on the group level, while observing more pronounced differences between EFO conditions on the subject level. Additionally, we demonstrated a trend towards higher MEPs with increasing number of trials.

### 4.1. Effect of trial number

Both in our regression model as well as in our correlation analyses, we observed a robust group-level trend towards higher MEP amplitudes across trial blocks. Importantly, this was also present in the SHAM group, where no effective NM via cTBS should have taken place. These findings may replicate findings from a recent study by Boucher et al., who have noted a similar pattern of MEP facilitation following sham TBS ^5^, as well as those of a study by Pellicciari et al., who observed increasing MEP amplitudes with cumulative single-pulse TMS application, irrespective of either fixed or random inter-stimulus intervals ^39^. Given that our findings are in agreement ^5, 39^, the finding that modulation of MEPs occurs even when individual stimuli are repeated at a rather large and randomized inter-stimulus interval is emphasized (5.4 - 6.6 s in our case).

While there is speculation as to the neurophysiological basis of this phenomenon ^39^, there is currently no conclusive explanation for it. Studies into spiking behavior of neurons following single pulses of TMS indicate changes in activity confined to 100-500 ms, which would imply that a stimulus following after this period should not lead to cumulative changes ^40, 41^. However, ours and the above-mentioned findings ^5, 39^ indicate that some form of longitudinal change must take place to explain the observed facilitation of MEPs. While outside of the scope of this study, variables that might plausibly change in the time range relevant to our inter-stimulation intervals could be found in metabolic parameters such as cerebral blood flow (CBF) or parenchymal oxygen content ^42, 43^. In this context, Allen et al. demonstrated that oxygen levels sharply increased in the first 10 s following a TMS pulse ^43^. Relatedly, Mesquita et al. demonstrated increasing CBF during the first 15-20 minutes of low-frequency repetitive TMS ^42^. If cumulative changes on this level instead of the single-cell level were hypothesized to contribute to the observed MEP amplitude facilitation, a combination of TMS paradigms with e.g. perfusion imaging could be attempted in the future to investigate said hypothesis. Specifically, it would be interesting whether a plateau in e. g. CBF changes as the one observed by Mesquita et al. were corresponding to a plateau in MEP facilitation ^42^. Additionally, adding CBF dynamics to e-field modeling could potentially improve e-field estimations across longer TMS experiments. As a more immediate takeaway, these results emphasize the need to consider trial-related variance of MEPs in any study employing MEPs as a relevant readout, including but not limited to the NM context.

### 4.2. Effect of EFO during cTBS

According to our results, EFO during cTBS emerged as a relevant parameter influencing MEP amplitudes in our mixed effects regression analysis. Specifically, on the group level, application of cTBS in condition 90 was predicted to lead to approximately 14% increased MEP amplitudes compared to cTBS in OPT (Table 2). The finding that EFO is a variable of interest in TMS for NM is plausible in light of many recent studies ^18, 19, 23, 30^ ^23^. Specifically, EFO not only influences the distribution and magnitude of the electric field acting upon the cortex ^18, 19, 30^, but also partially determines which neuron populations are preferentially stimulated ^23^.

Considering that cTBS affects MEPs via indirect transsynaptic conduction (e.g., in the form of I-wave suppression) ^24^, it stands to reason that EFO tailored to target specific neuron subpopulations could optimize NM results. This perspective could also contribute to explaining the substantial variability observed in aftereffects of cTBS ^24^, since individual differences in gyral geometry could lead to variance in optimal EFO for NM application.

While the exact origin of MEPs is complex to determine due to the multitude of neurons simultaneously affected by a TMS pulse, the directional sensitivity of MEP amplitudes is well established ^44^. If not specifically subject to investigation, the optimal orientation for MEP elicitation is often presumed to be perpendicular to the precentral gyrus, despite evidence for deviations from this heuristic principle ^31^. For NM, little evidence for optimal EFO exists, despite optimization studies for other parameters such as intensity or number of stimuli ^45, 46^. Our results can however be interpreted in a way suggesting that optimal EFO for MEP generation may not always be optimal for NM protocols at the same site.

Whether one considers our findings as pointing towards the 90 condition being superior to the OPT condition on the group-level remains open for debate; on the one hand, the 90 condition led to higher MEP facilitation compared to OPT; on the other hand, the traditionally expected outcome of cTBS would be MEP suppression ^24, 26^, which would render OPT the condition less deviating from the expected result. Additionally, our group-level results fail to properly capture the significant heterogeneities observed on the single-subject level, which indicates that a “one-size-fits-all” approach in optimizing EFO should be avoided. From a more application-oriented perspective, our findings serve to yield evidence that EFO is generally an optimizable parameter in NM.

### 4.3. Implications for use cases of NM

Considering the large available variety and increasing application of NM protocols, our results imply a need to conduct further research into the EFO parameter to better understand its potential impact on therapeutic TMS applications. In the specific case of depression therapy for example, one of the most well-established applications of NM by TMS, stimulation frequency^2^, target selection for stimulation ^7^, tailoring of treatment to brain state ^47^, or individual functional connectivity ^12^ have been subjects of investigation, with a general trend towards better effects for more individualized stimulation approaches ^7^. Comparatively little work has been performed in optimizing EFO for clinical use cases of NM ^1, 48^. One of the few studies that have specifically tested EFO in a clinical context was published in 2008, reporting differential outcomes to TMS treatment of chronic pain based on EFO during the applied TMS protocol ^48^. It stands to reason that optimization of EFO may be a promising avenue in dealing with heterogeneities in treatment responses. Considering the strong inter-individual variance in response to NM at different EFO, it may even be ideal to conduct a diagnostic process on a case-by-case basis to account for as many idiosyncrasies as possible and establish optimal stimulation parameters for each subject. Investigations into this topic could profit from the recent advances made in subject-specific e-field tailoring to take into account individual gyral geometries ^18, 19, 23^.

### 4.4. Limitations

First, our assumptions in the power analysis prior to recruiting were not accurate when compared to the final data, with a notable underestimation of MEP variability. Nonetheless, our regression modeling approach enhanced by bootstrapping implied robust effects of our explanatory variables. Second, the definition of OPT, 90, and SHAM was conducted based on the directional MEP data from the first session, and subsequently kept invariant (Figure 1). We can therefore not rule out that changes in directional sensitivity between sessions may have confounded our results. Third, the directional testing before the actual MEP experiment may have influenced our results, since 140 stimuli were applied to the motor hotspot at a stimulation intensity close to the subsequently determined rMT. However, this step was kept constant between sessions to ensure that if a bias was related to this, it would be stable across sessions. Relatedly, we did not perform subject-specific e-field modeling, but based our assessment of the optimal direction for MEP generation strictly on the empirical data. Fourth, the effects based on differences in EFO during NM are rather small, with an approximate 14% difference in MEP amplitudes between the two active conditions. It is unclear whether this difference would translate into meaningful clinical differences, and whether this finding would be generalizable to non-motor areas. Fifth, while we recorded EMG data from three muscles, all recordings were conducted during stimulation of only the APB motor hotspot. Other cortical locations may have been preferable for FDI and ADM assessments.

## 5. Conclusions

We investigated cTBS-related MEP modulation in healthy subjects across two verum conditions differing in EFO and one sham condition. We observed significant changes in MEP amplitudes based on EFO during NM, as well as a trend towards higher MEP amplitudes across trial blocks. Our results may yield evidence for the relevance of EFO during TMS-based NM protocols and may motivate further research into subject-tailored applications of NM.

## Data Availability

All data produced in the present study are available upon reasonable request to the authors

## 6. Acknowledgements

We thank the participants for taking part in our study.

## Abbreviations

3D: Three-dimensional
ADM: Adductor digiti minimi
ANOVA: Analysis of variance
APB: Abductor pollicis brevis
cTBS: Continuous theta burst stimulation
EEG: Electroencephalography
EFO: E-field orientation
EMG: Electromyography
FDI: First dorsal interosseus
fMRI: Functional magnetic resonance imaging
MEP: Motor-evoked potential
MSO: Maximum stimulator output
NM: Neuromodulation
rMT: Resting motor threshold
TMS: Transcranial magnetic stimulation

