## Supplementary Material for "Number of Trials and E-Field Orientation during Continuous Theta Burst Stimulation May Impact Modulation of Motor-Evoked Potentials"

Supplementary Figure 1:
Plot of Residuals during Linear Mixed Effects Regression Modeling of Untransformed MEP Data

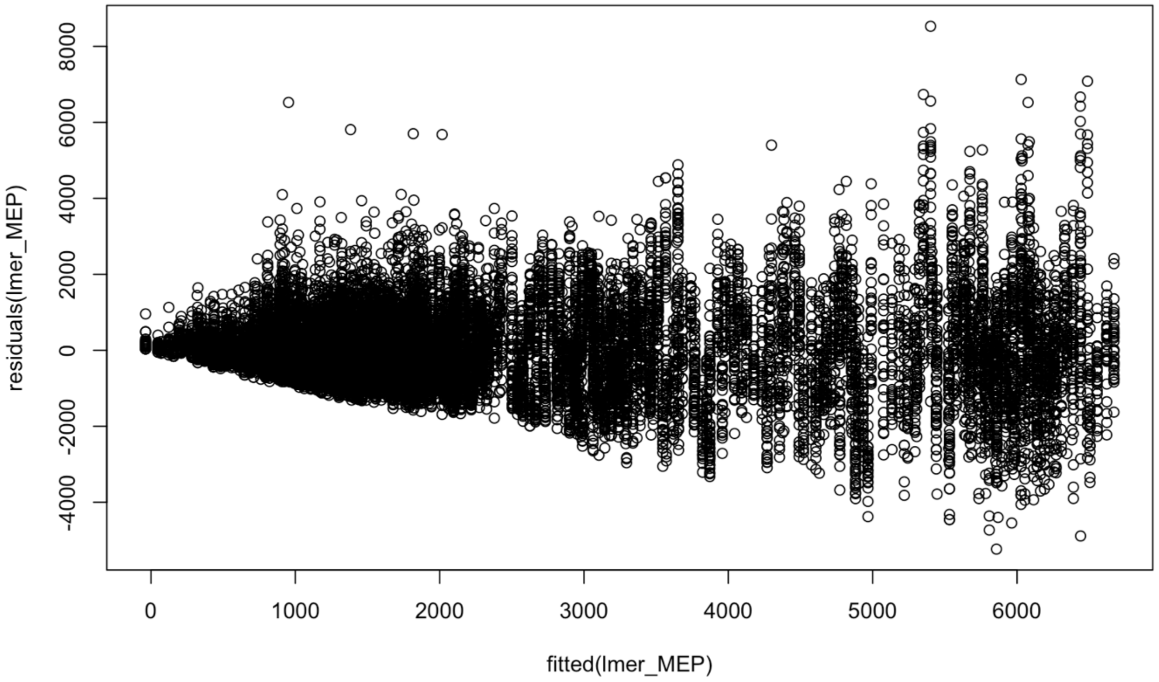


Note the funnel-like shape of the residuals, indicating that higher MEP amplitudes were subject to worse fit. This was interpreted to result from heteroscedasticity inherent in the data, and motivated the log-transform of the MEPs.

Supplementary Figures 2-18:

Additional subject-specific MEP amplitude distributions split by muscle and TBS condition across trial blocks. Note missing values as noted in the beginning of section 3 in the main manuscript. As specified in the main manuscript, subject 8 was excluded from further sessions due to the described incidental finding.

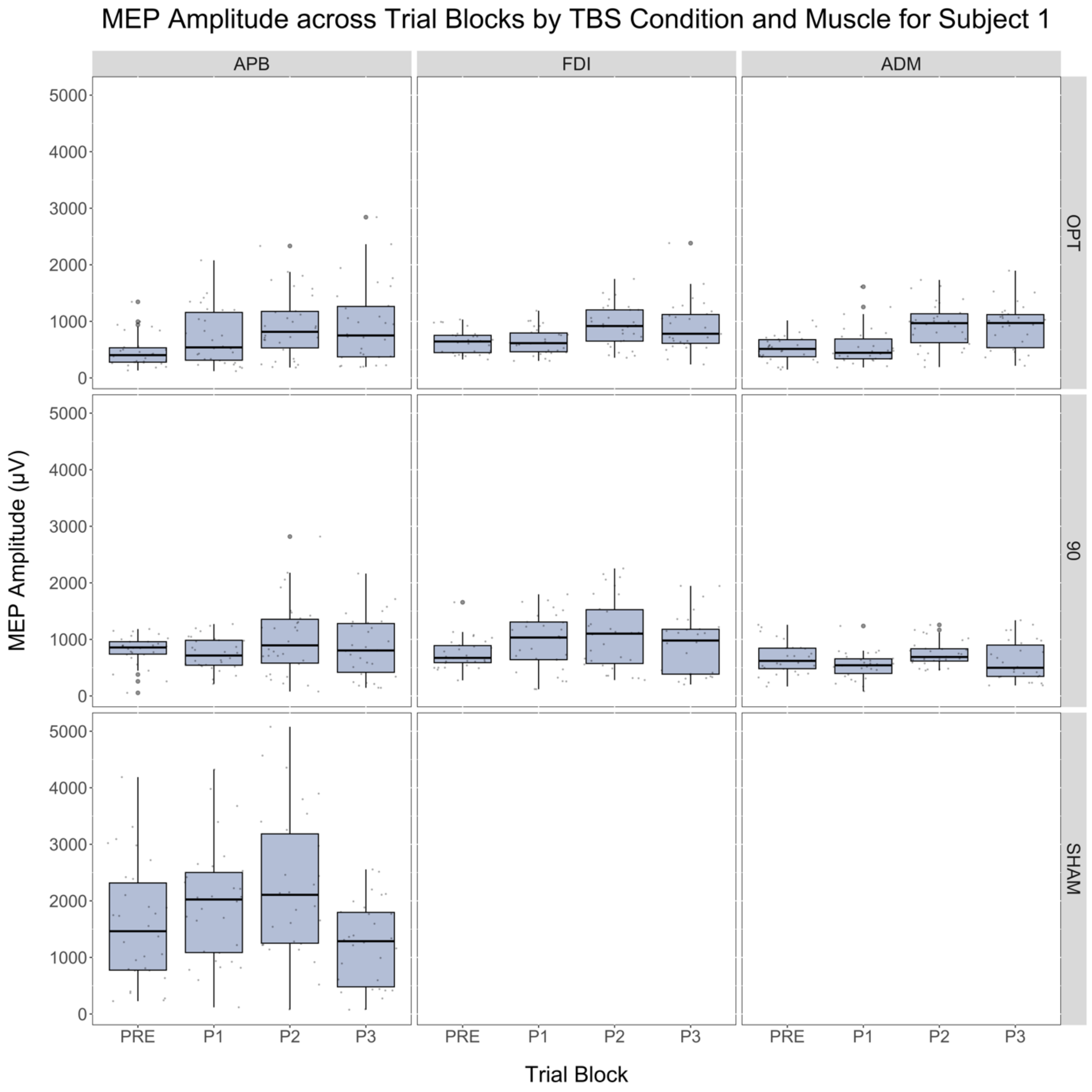
MEP Amplitude across Trial Blocks by TBS Condition and Muscle for Subject 1
APB
FDI
ADM
OPT
90
SHAM
MEP Amplitude (μV)
0
1000
2000
3000
4000
5000
PRE
P1
P2
P3
Trial Block

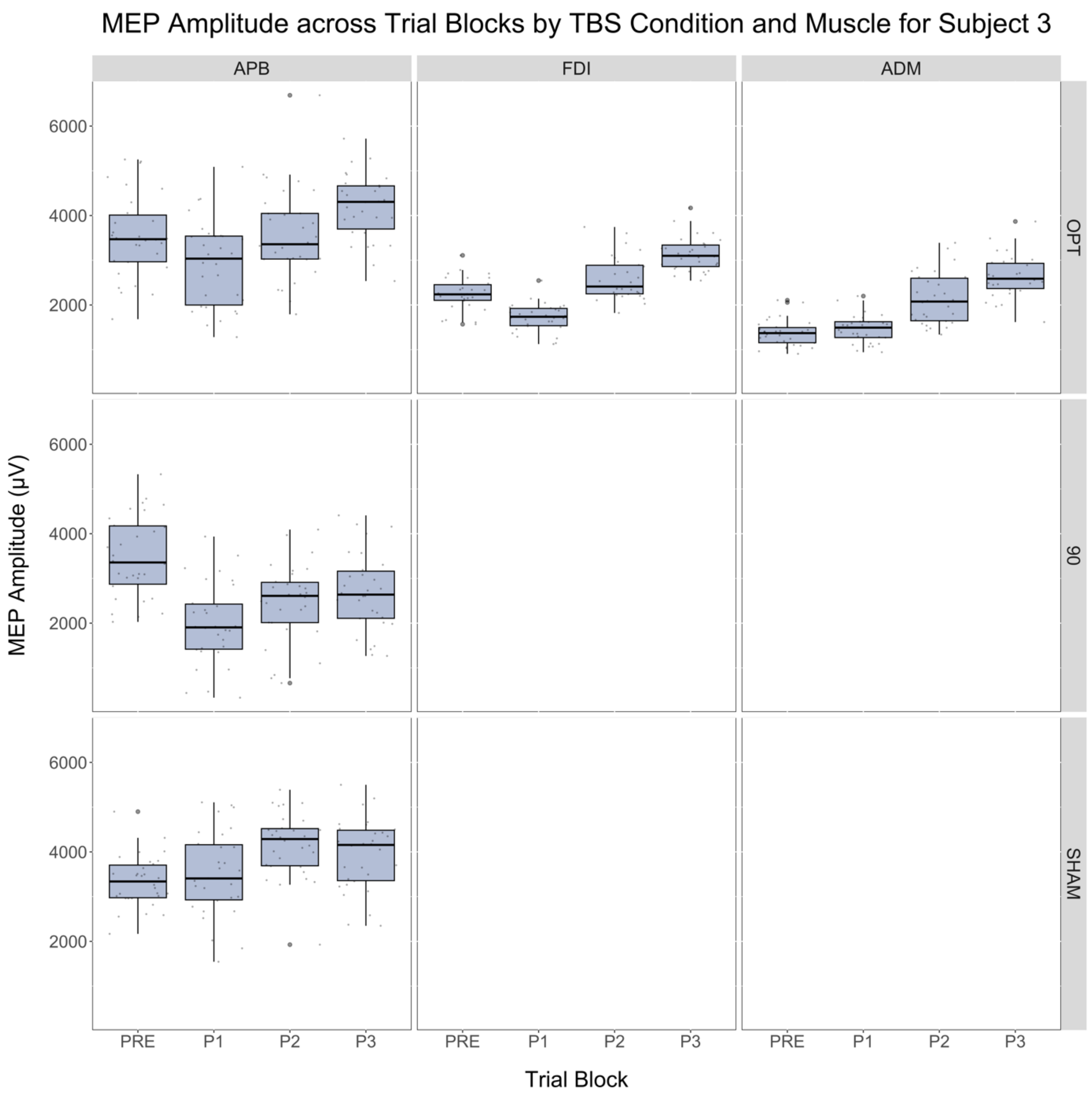

MEP Amplitude across Trial Blocks by TBS Condition and Muscle for Subject 3
APB
FDI
ADM
OPT
90
SHAM
MEP Amplitude (µV)
6000
4000
2000
PRE
P1
P2
P3
Trial Block

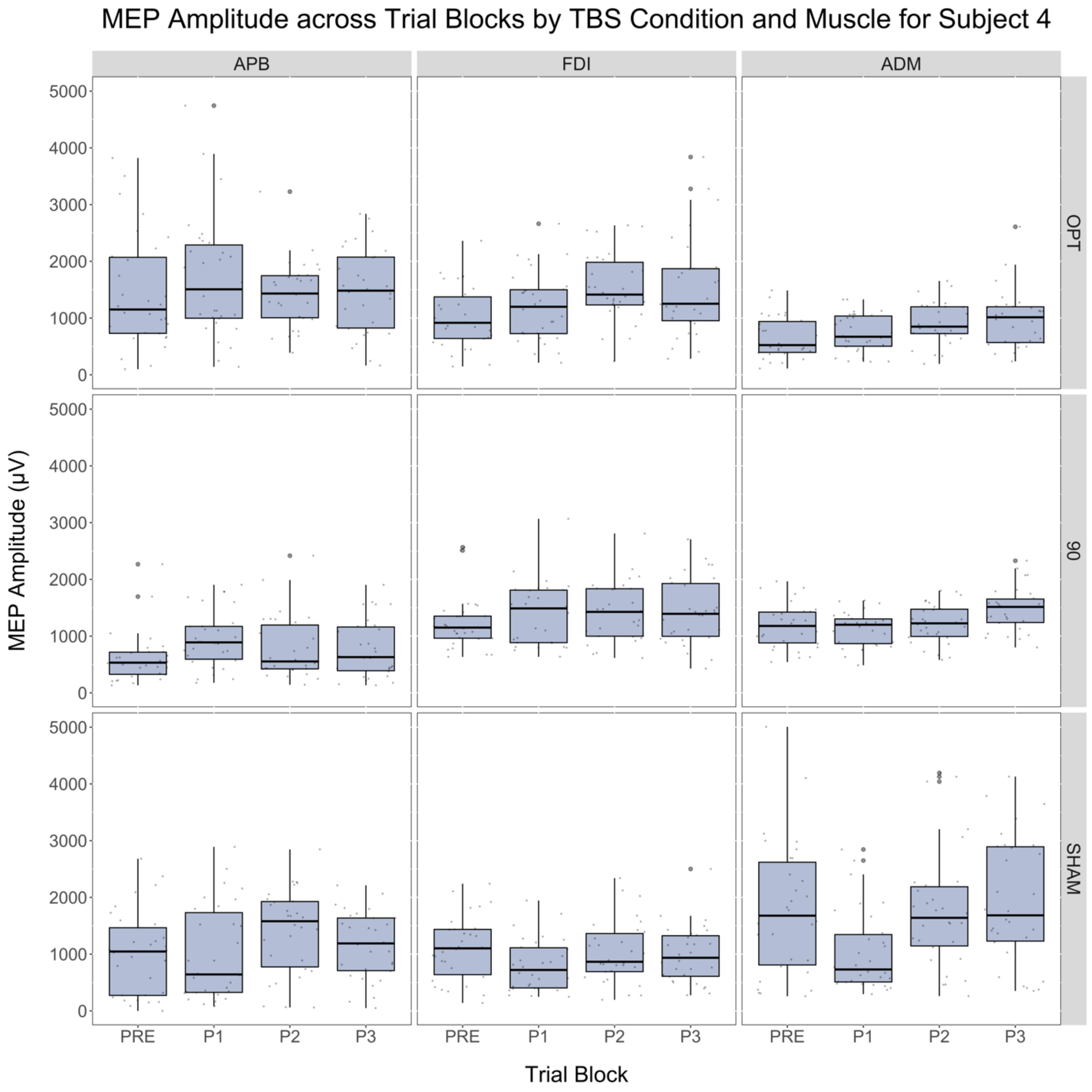

MEP Amplitude across Trial Blocks by TBS Condition and Muscle for Subject 4
APB
FDI
ADM
OPT
90
SHAM
MEP Amplitude (µV)
0
1000
2000
3000
4000
5000
PRE
P1
P2
P3
Trial Block

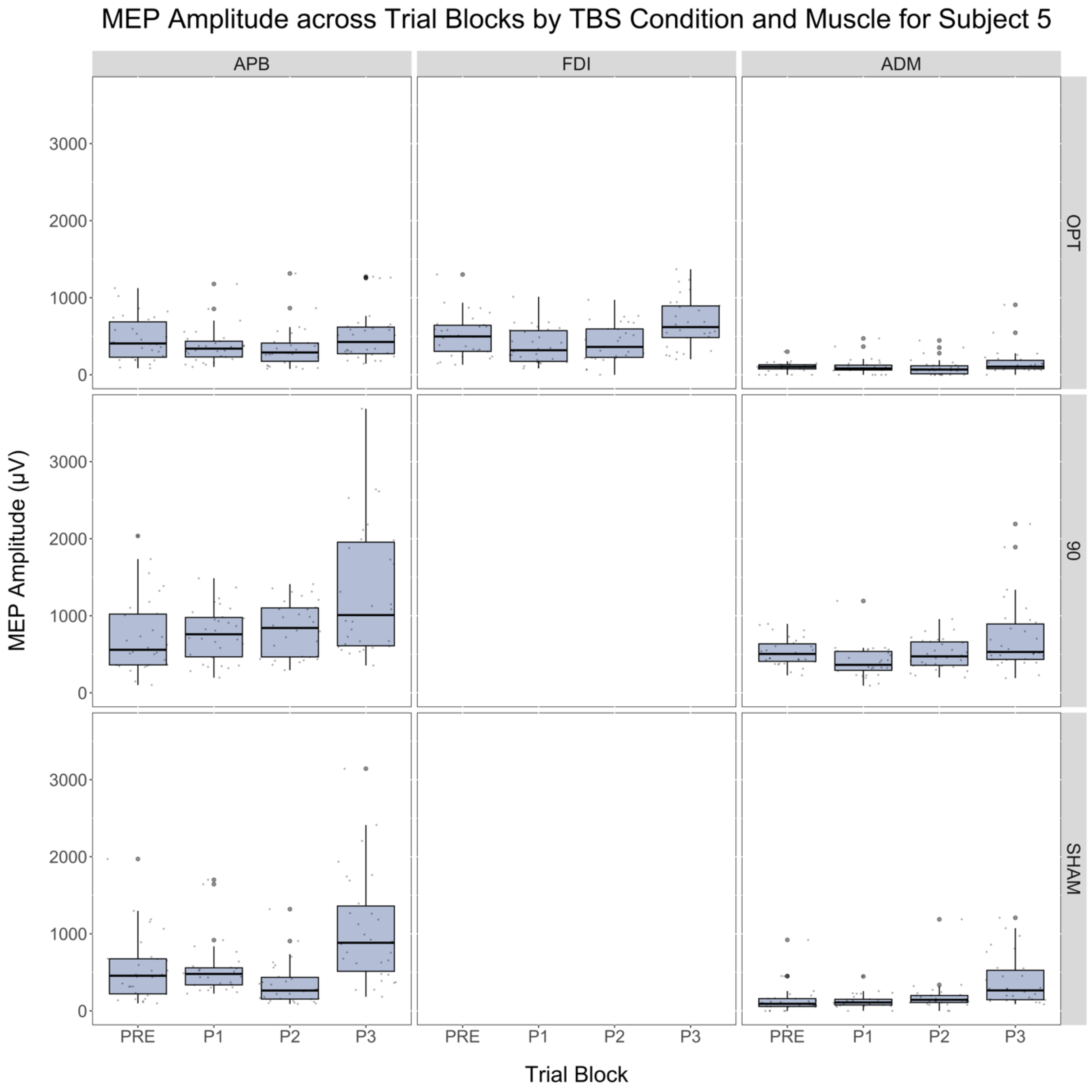

MEP Amplitude across Trial Blocks by TBS Condition and Muscle for Subject 5
APB
FDI
ADM
OPT
90
SHAM
MEP Amplitude (µV)
0
1000
2000
3000
PRE
P1
P2
P3
Trial Block

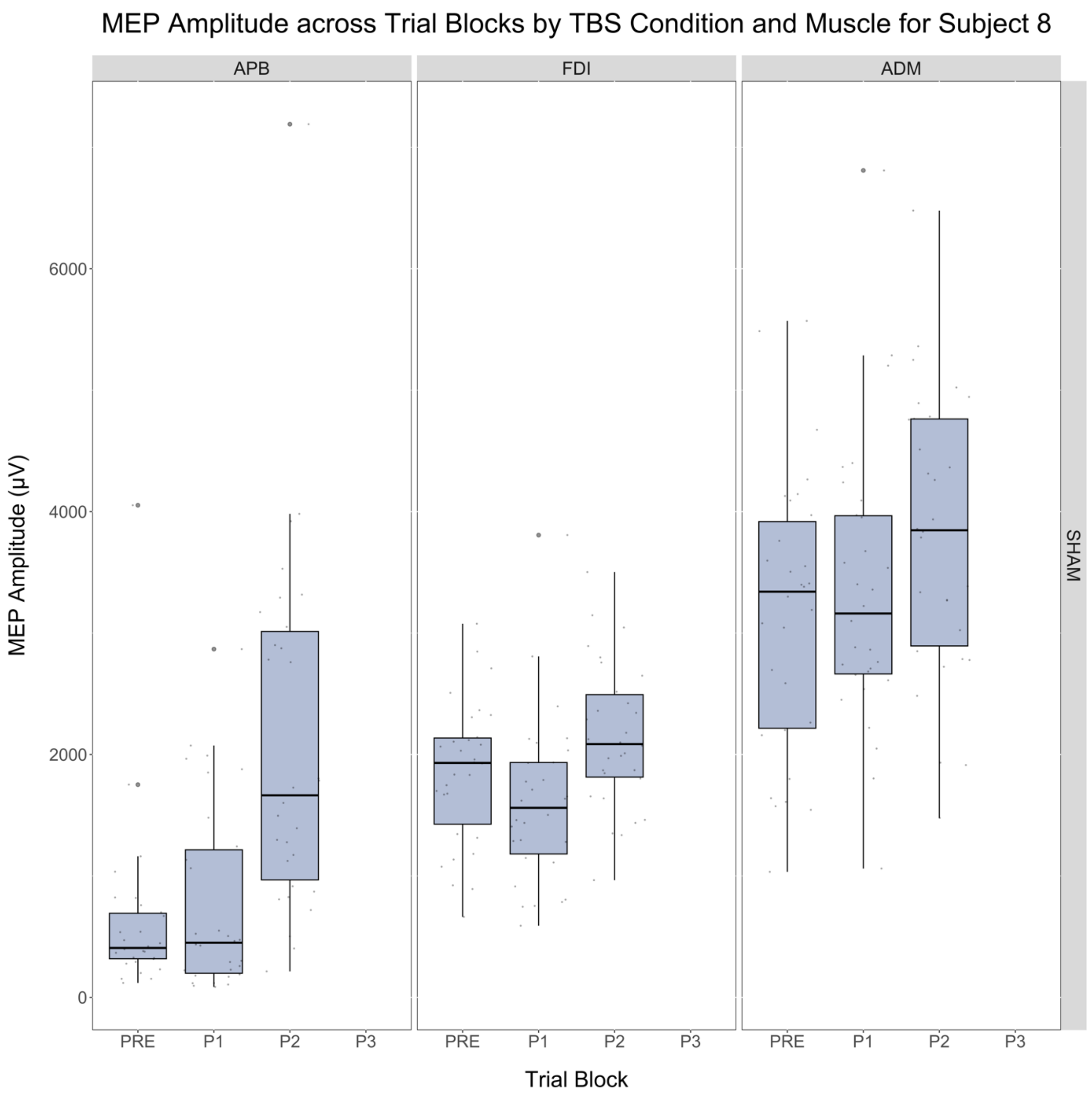
MEP Amplitude across Trial Blocks by TBS Condition and Muscle for Subject 8
APB
FDI
ADM
SHAM
MEP Amplitude (μV)
0
2000
4000
6000
PRE
P1
P2
P3
Trial Block

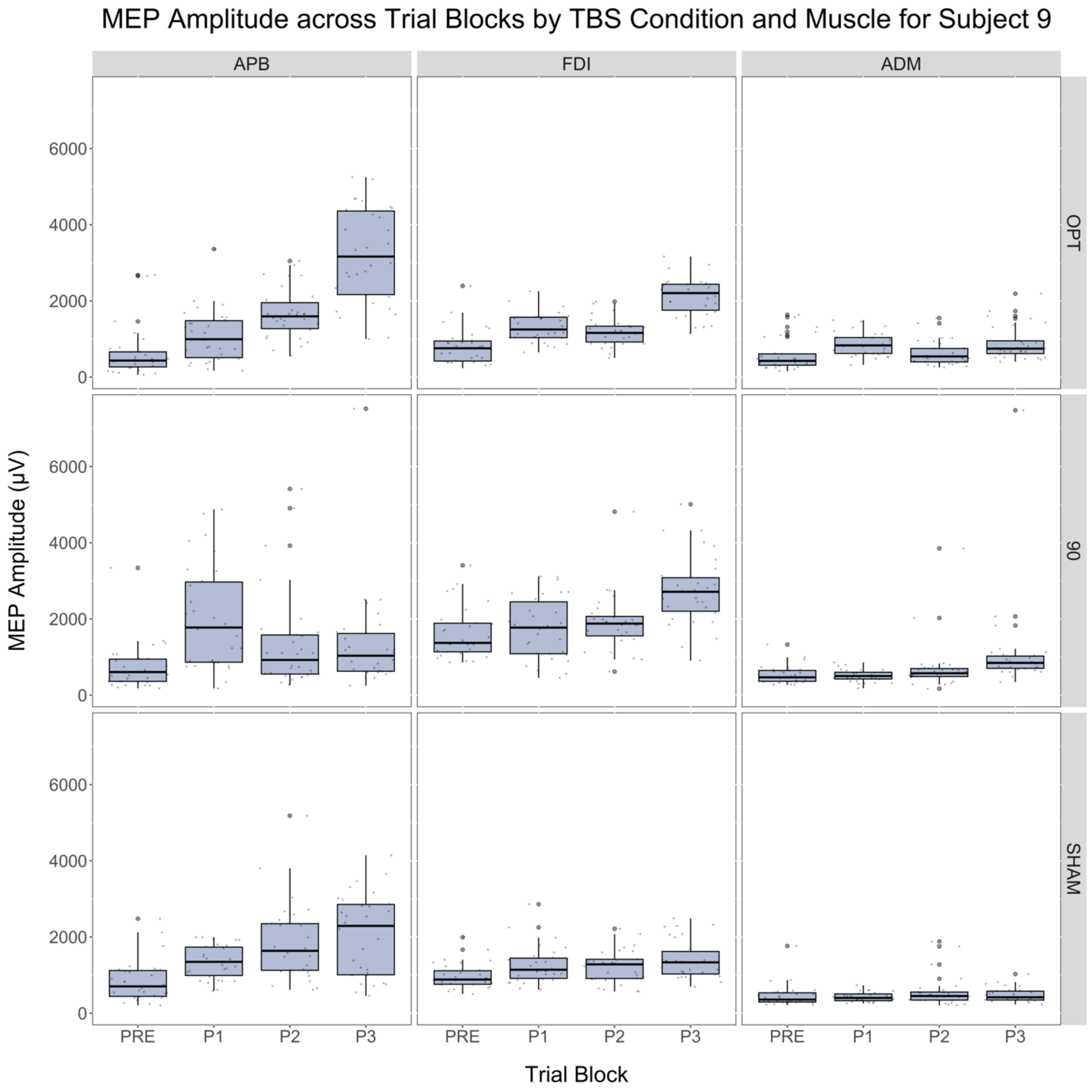

MEP Amplitude across Trial Blocks by TBS Condition and Muscle for Subject 9
APB
FDI
ADM
OPT
90
SHAM
MEP Amplitude (μV)
0
2000
4000
6000
PRE
P1
P2
P3
Trial Block

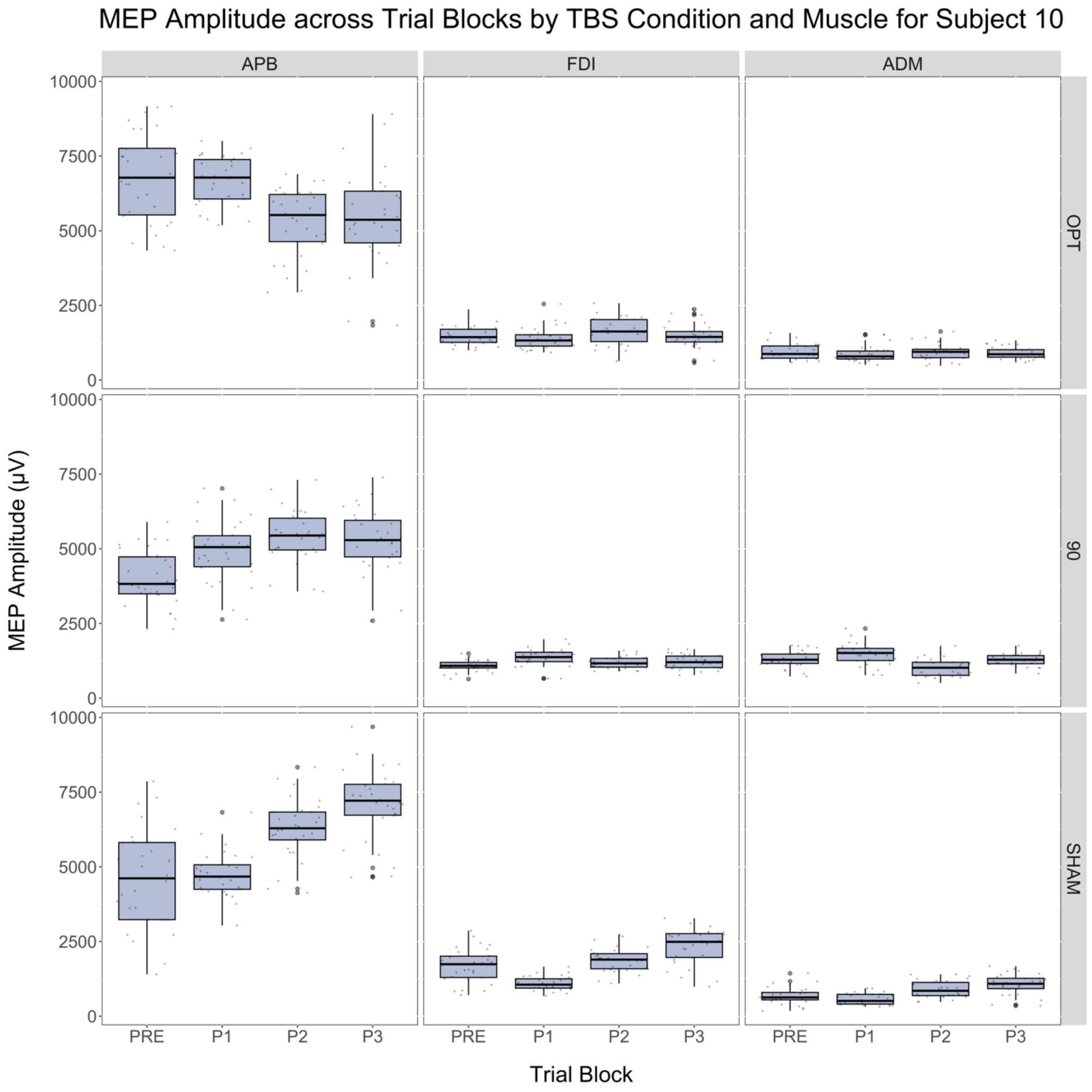
MEP Amplitude across Trial Blocks by TBS Condition and Muscle for Subject 10
APB
FDI
ADM
OPT
90
SHAM
MEP Amplitude (μV)
0
2500
5000
7500
10000
PRE
P1
P2
P3
Trial Block

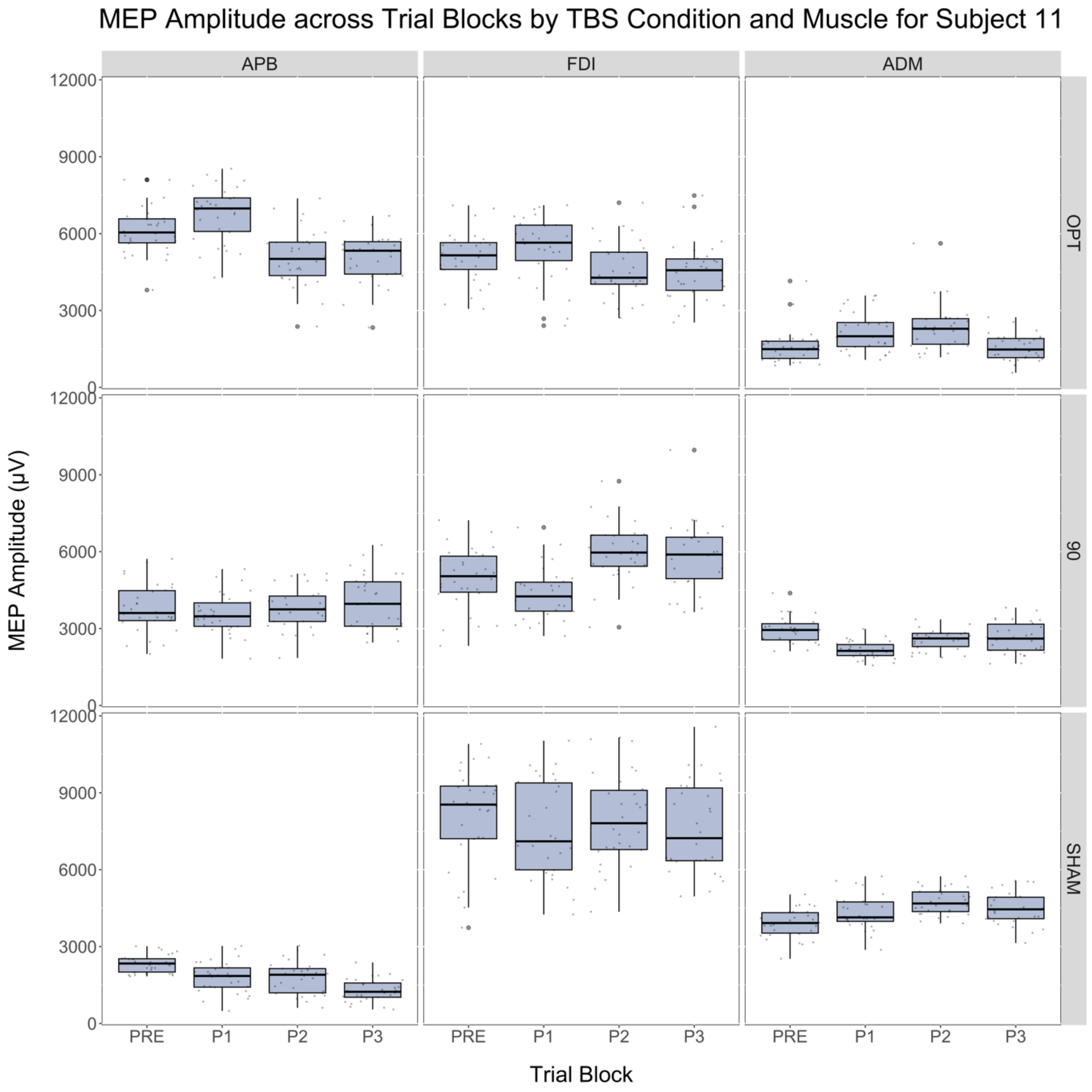
MEP Amplitude across Trial Blocks by TBS Condition and Muscle for Subject 11
APB
FDI
ADM
OPT
90
SHAM
MEP Amplitude (μV)
12000
9000
6000
3000
0
PRE
P1
P2
P3
Trial Block

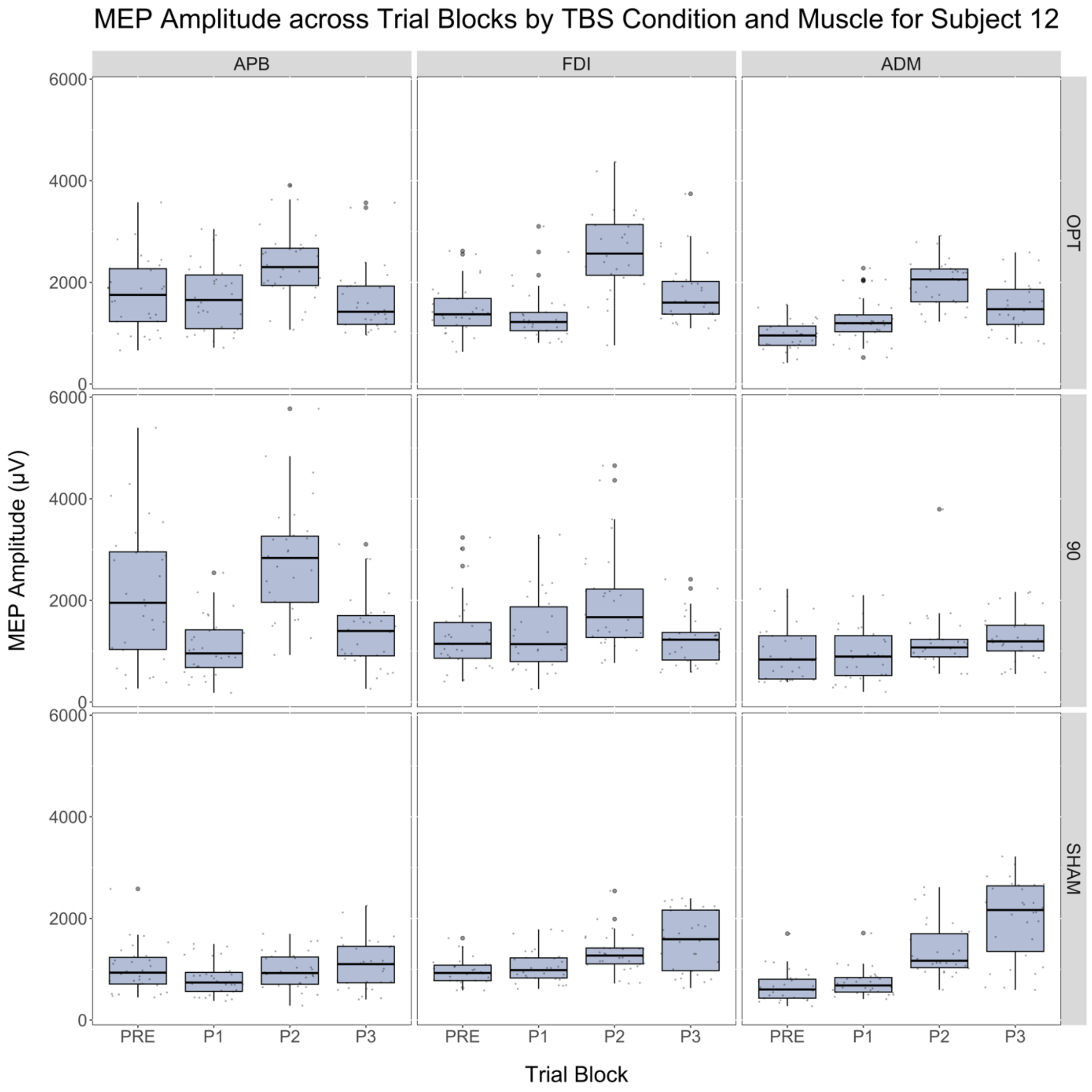
MEP Amplitude across Trial Blocks by TBS Condition and Muscle for Subject 12
APB
FDI
ADM
OPT
90
SHAM
MEP Amplitude (µV)
0
2000
4000
6000
PRE
P1
P2
P3
Trial Block

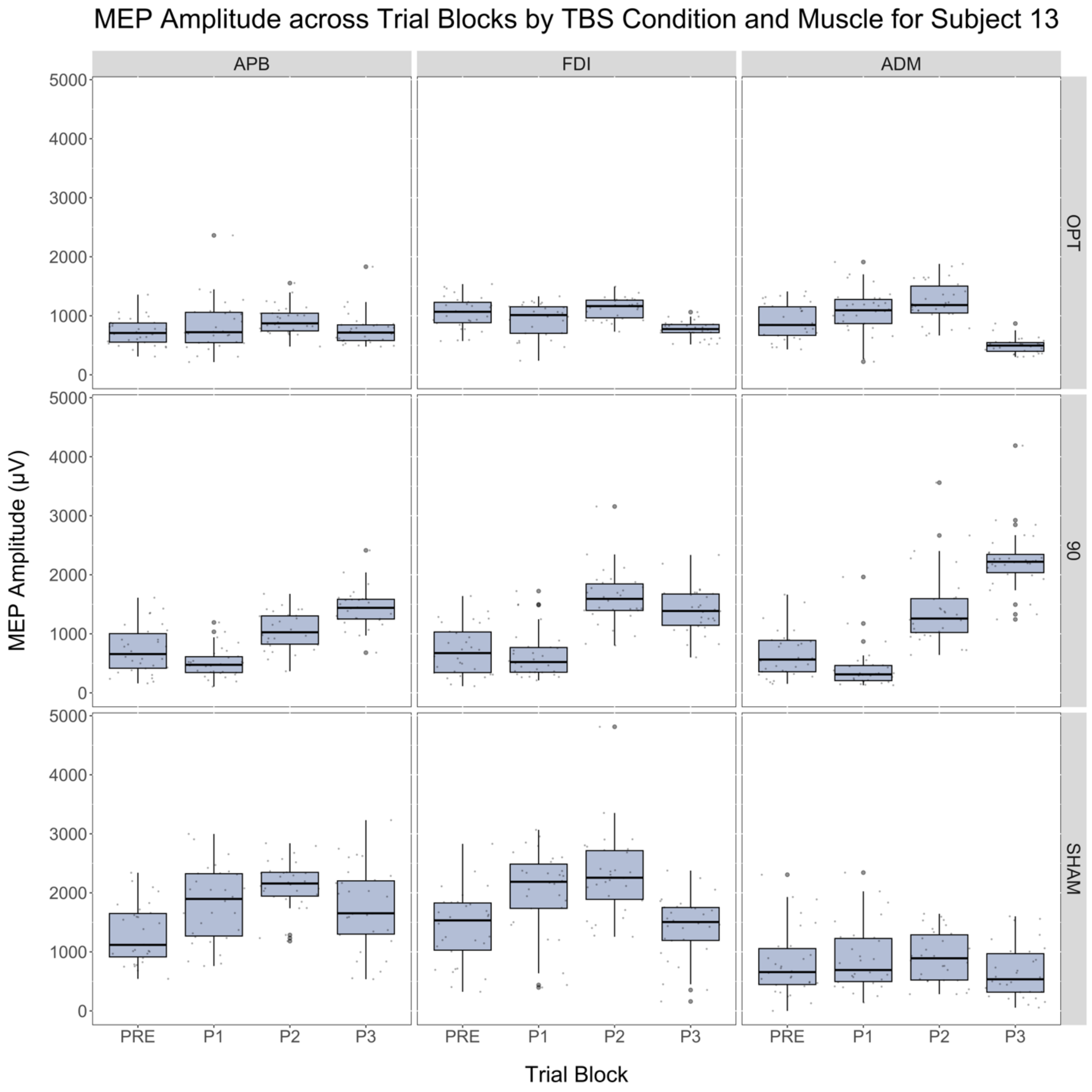

MEP Amplitude across Trial Blocks by TBS Condition and Muscle for Subject 13
APB
FDI
ADM
OPT
90
SHAM
MEP Amplitude (μV)
0
1000
2000
3000
4000
5000
PRE
P1
P2
P3
Trial Block

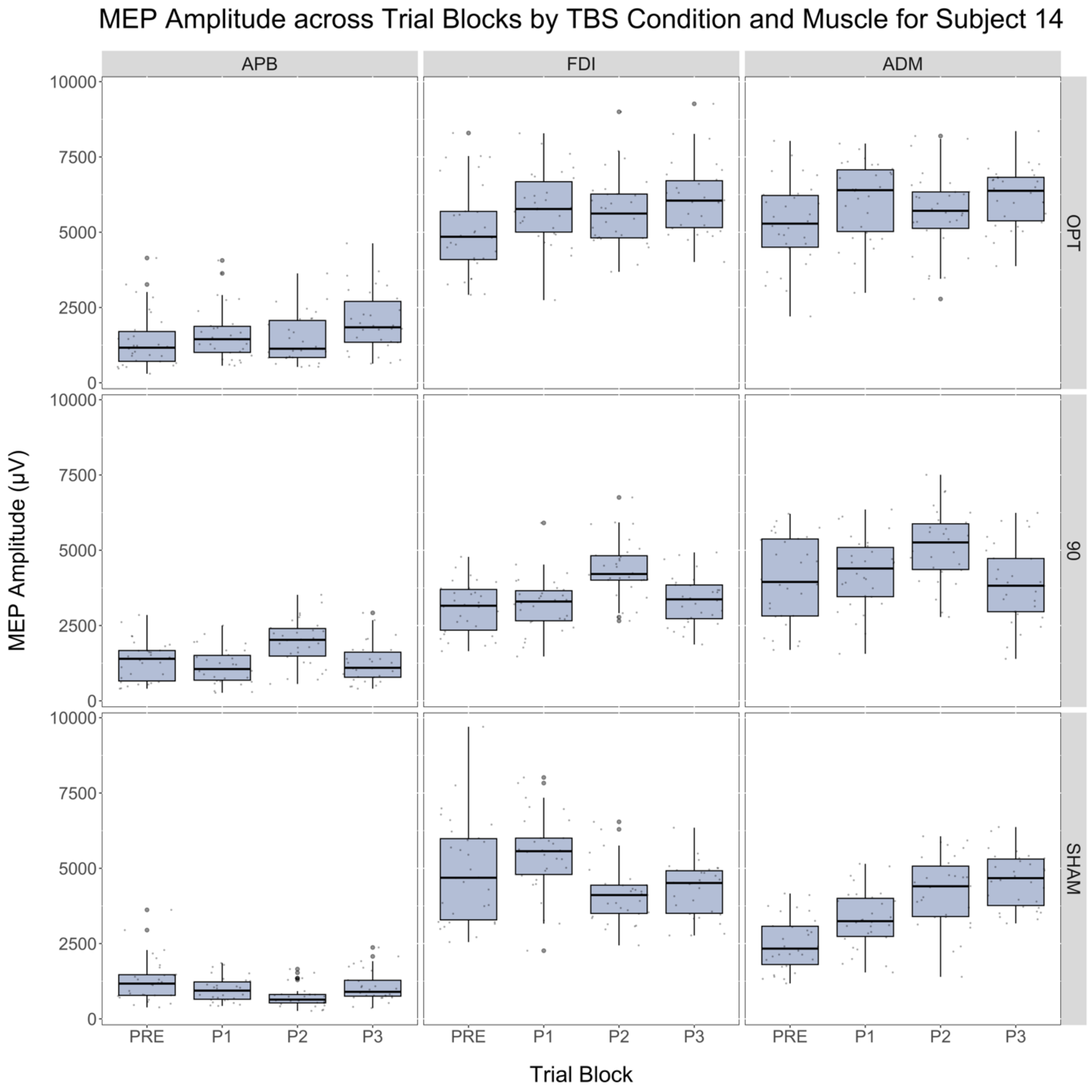
MEP Amplitude across Trial Blocks by TBS Condition and Muscle for Subject 14
APB
FDI
ADM
OPT
90
SHAM
MEP Amplitude (μV)
10000
7500
5000
2500
0
PRE
P1
P2
P3
Trial Block

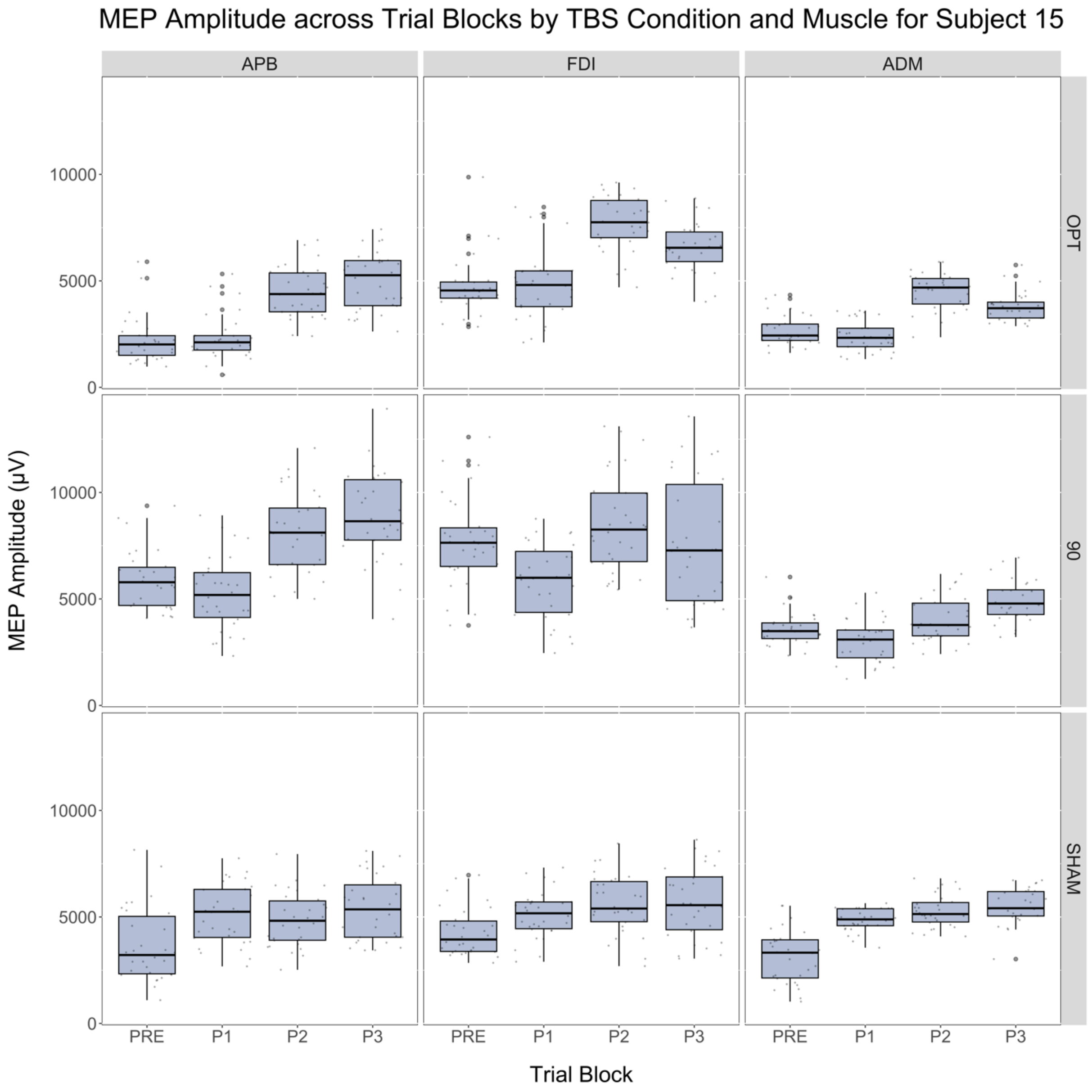
MEP Amplitude across Trial Blocks by TBS Condition and Muscle for Subject 15
APB
FDI
ADM
OPT
90
SHAM
MEP Amplitude (µV)
0
5000
10000
PRE
P1
P2
P3
Trial Block

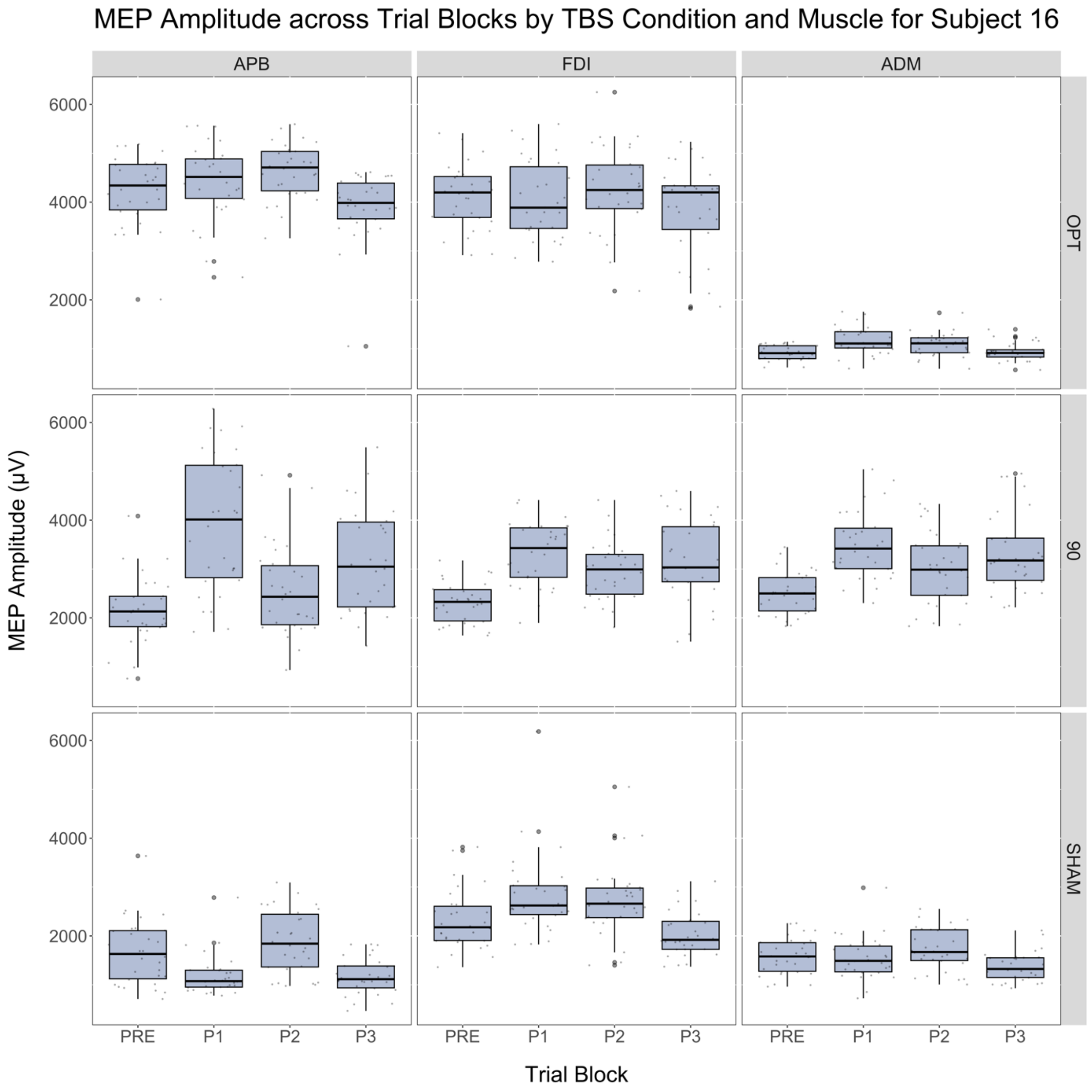

MEP Amplitude across Trial Blocks by TBS Condition and Muscle for Subject 16
APB
FDI
ADM
OPT
90
SHAM
MEP Amplitude (μV)
6000
4000
2000
PRE
P1
P2
P3
Trial Block

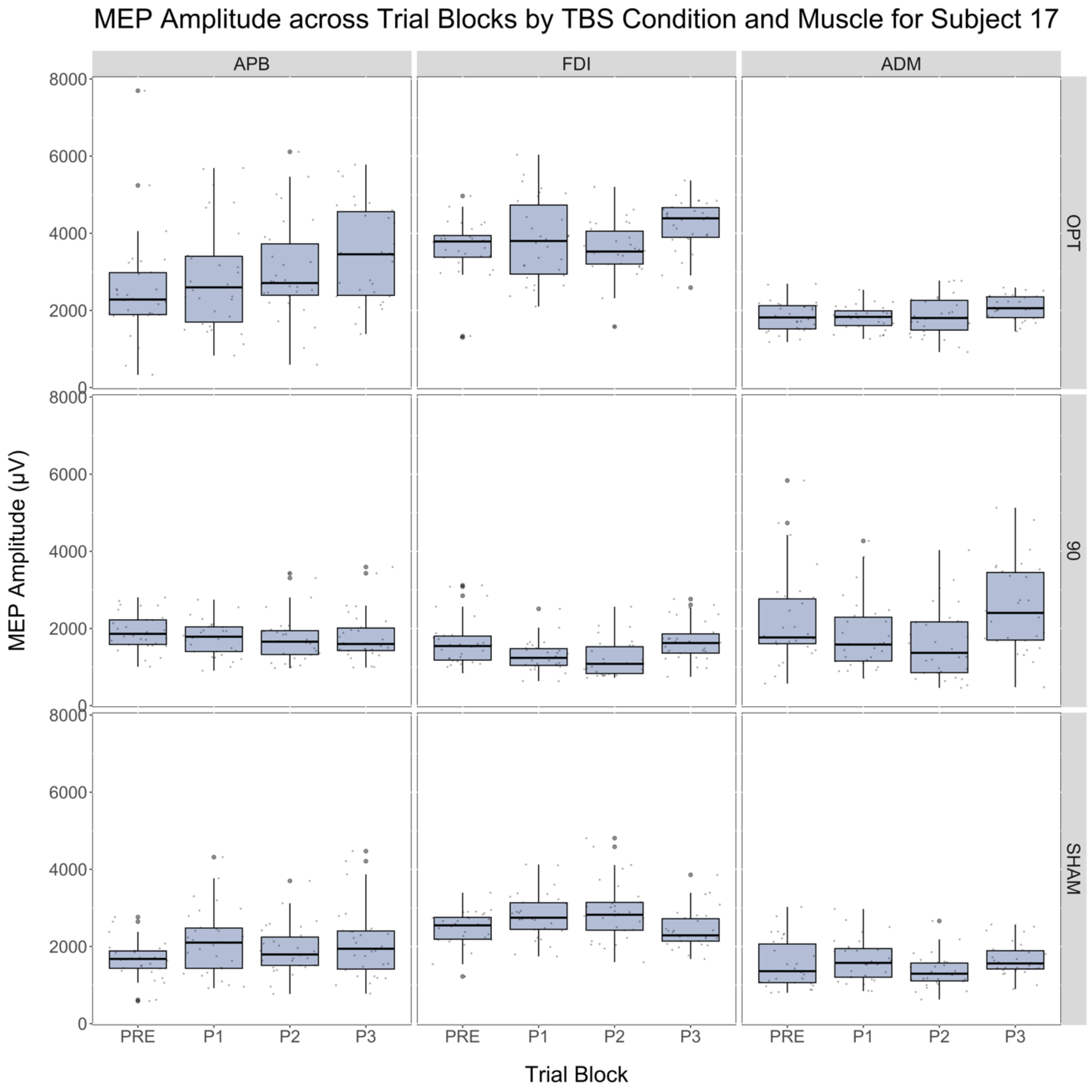

MEP Amplitude across Trial Blocks by TBS Condition and Muscle for Subject 17
APB
FDI
ADM
OPT
90
SHAM
MEP Amplitude (µV)
8000
6000
4000
2000
0
PRE
P1
P2
P3
Trial Block

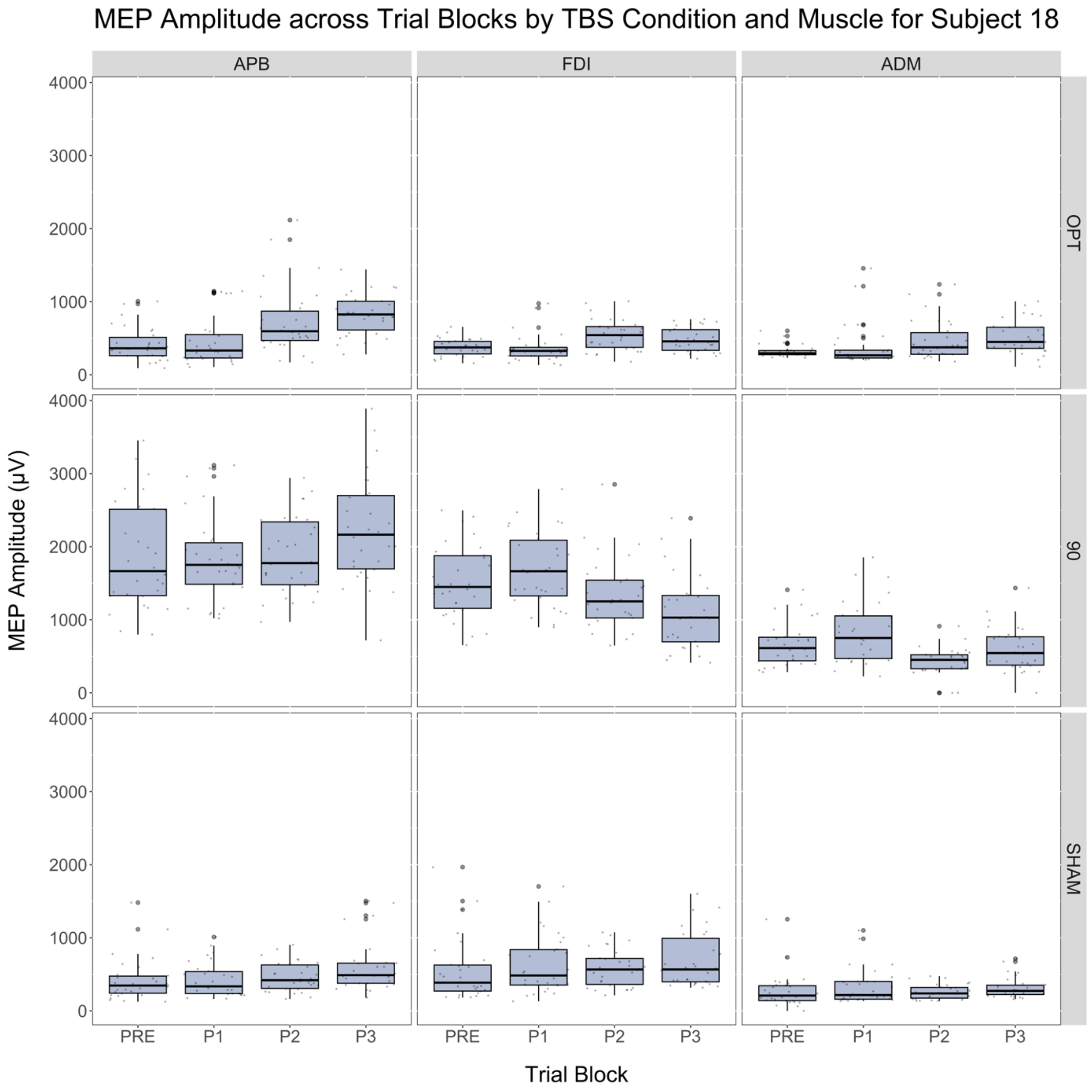
MEP Amplitude across Trial Blocks by TBS Condition and Muscle for Subject 18
APB
FDI
ADM
OPT
90
SHAM
MEP Amplitude (μV)
4000
3000
2000
1000
0
PRE
P1
P2
P3
Trial Block

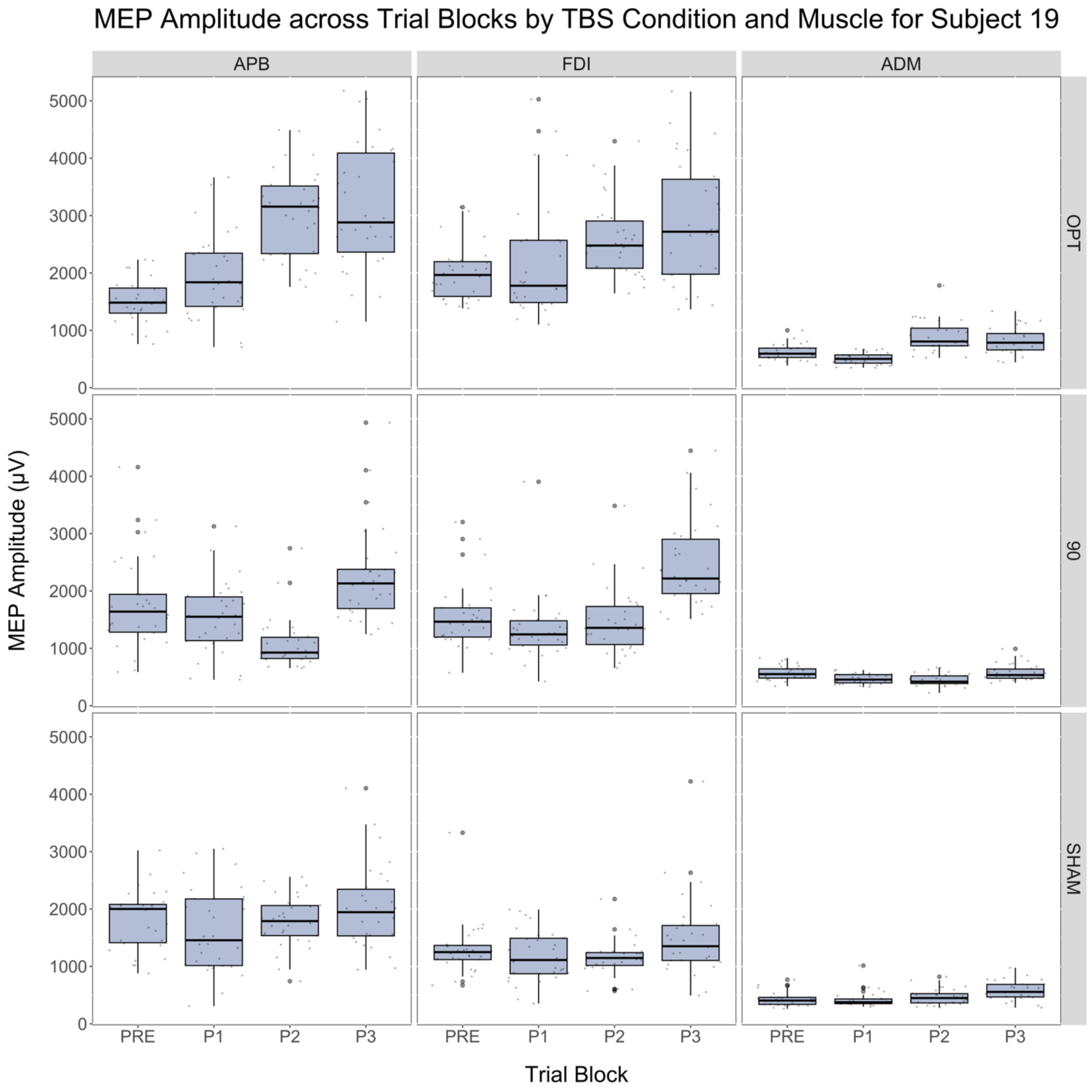
MEP Amplitude across Trial Blocks by TBS Condition and Muscle for Subject 19
APB
FDI
ADM
OPT
90
SHAM
MEP Amplitude (µV)
0
1000
2000
3000
4000
5000
PRE
P1
P2
P3
Trial Block

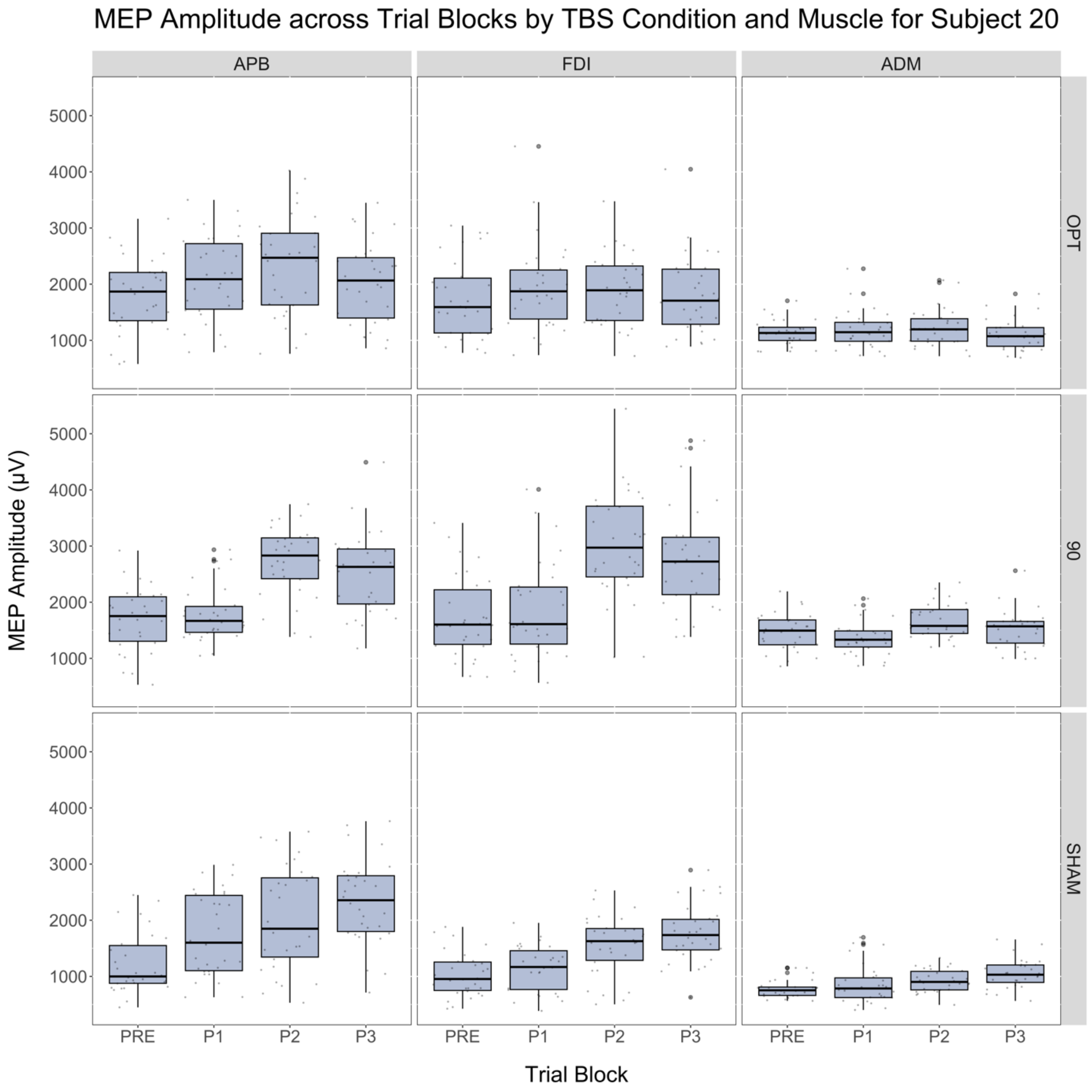

MEP Amplitude across Trial Blocks by TBS Condition and Muscle for Subject 20
APB
FDI
ADM
OPT
90
SHAM
MEP Amplitude (μV)
1000
2000
3000
4000
5000
PRE
P1
P2
P3
Trial Block
